# Serum proteomic profiling of physical activity reveals CD300LG as a novel exerkine with a potential causal link to glucose homeostasis

**DOI:** 10.1101/2024.02.10.24302626

**Authors:** Sindre Lee-Ødegård, Marit Hjorth, Thomas Olsen, Gunn-Helen Moen, Emily Daubney, David M Evans, Andrea Hevener, Aldons Jake Lusis, Mingqi Zhou, Marcus Michael Seldin, Hooman Allayee, James R. Hilser, Jonas Krag Viken, Hanne L. Gulseth, Frode Norheim, Christian A. Drevon, Kåre I. Birkeland

## Abstract

**Background:** Physical activity has been associated with preventing the development of type 2 diabetes and atherosclerotic cardiovascular disease. However, our understanding of the precise molecular mechanisms underlying these effects remains incomplete and good biomarkers to objectively assess physical activity are lacking.

**Methods:** We analyzed 3072 serum proteins in 26 men, normal weight or overweight, undergoing 12 weeks of a combined strength and endurance exercise intervention. We estimated insulin sensitivity with hyperinsulinemic euglycemic clamp, maximum oxygen uptake, muscle strength, and used MRI/MRS to evaluate body composition and organ fat depots. Muscle and subcutaneous adipose tissue biopsies were used for mRNA sequencing. Additional association analyses were performed in samples from up to 47,747 individuals in the UK Biobank, as well as using 2-sample Mendelian randomization and mice models.

**Results:** Following 12 weeks of exercise intervention, we observed significant changes in 283 serum proteins. Notably, 66 of these proteins were elevated in overweight men and positively associated with liver fat before the exercise regimen, but were normalized after exercise. Furthermore, for 19.7% and 12.1% of the exercise-responsive proteins, corresponding changes in mRNA expression levels in muscle and fat, respectively, were shown. The protein CD300LG displayed consistent alterations in blood, muscle, and fat. Serum CD300LG exhibited positive associations with insulin sensitivity, and to angiogenesis-related gene expression in both muscle and fat. Furthermore, serum CD300LG was positively associated with physical activity and negatively associated with glucose levels in the UK Biobank. In this sample, the association between serum CD300LG and physical activity was significantly stronger in men than in women. Mendelian randomization analysis suggested potential causal relationships between levels of serum CD300LG and fasting glucose, 2-hour glucose after an oral glucose tolerance test, and HbA1c. Additionally, Cd300lg responded to exercise in a mouse model, and we observed signs of impaired glucose tolerance in male, but not female, *Cd300lg* knockout mice.

**Conclusion:** Our study identified several novel proteins in serum whose levels change in response to prolonged exercise and were significantly associated with body composition, liver fat, and glucose homeostasis. Serum CD300LG increased with physical activity and is a potential causal link to improved glucose levels. CD300LG may be a promising exercise biomarker and a therapeutic target in type 2 diabetes.

## Introduction

Physical activity is a cornerstone in the prevention and treatment of several chronic diseases like obesity, non-alcoholic fatty liver disease (NAFLD), atherosclerotic vascular disease, and type 2 diabetes mellitus ^1^. Both acute and long-term exercise may enhance insulin sensitivity and thereby improve glucose tolerance ^2^. Both resistance and endurance exercises enhance insulin sensitivity, although the most pronounced effect is observed when combining these training modalities ^3^.

Metabolic adaptations to exercise encompass intricate inter-organ communication facilitated by molecules referred to as exerkines ^4–7^. These exerkines are secreted from various tissues and include a variety of signal molecules released in response to acute and/or long-term exercise with endocrine, paracrine and/or autocrine functions ^4,5^. Although there has been considerable emphasis on exerkines originating from skeletal muscle (SkM) ^8,9^, it is also known that exerkines can originate from organs such as white ^6,10,11^ and brown adipose tissue ^12^ or the liver ^13^. The established *bona fide* exerkine, interleukin-6 (IL6), is released during muscle contractions, contributing to improved overall glucose homeostasis ^4,14^. In addition, a range of other exerkines are recognized, including IL7 ^15^, 12,13-diHOME ^12^, myonectin ^16^, myostatin ^17,18^, METRNL ^19^, CSF1 ^8^, decorin ^20^, SFRP4 ^10^, fetuin-A ^13,21^, and ANGPTL4 ^22,23^, among many others ^5^.

Extensive screening aimed at discovering novel exercise responsive blood proteins have faced considerable challenges, primarily due to the technical challenges in quantifying the blood proteome on a large scale. However, recent advances in multi-plex technology, such as the proximity extension assay (PEA), have made it possible to quantify more than 3000 proteins in blood samples more reliably than traditional untargeted mass spectrometry (https://olink.com/application/pea). Some recent studies have used other proteomic platforms, such as aptamer-based techniques (https://somalogic.com/somascan-platform/), to show that acute and long-term aerobic exercise affected several hundred serum proteins ^24–28^, but the downstream causal effects of such changes on clinical phenotypes are not known. Furthermore, no studies have used the PEA technology to identify exerkines potentially underlying the mechanisms through which long-term physical activity, including strength exercise, enhances glucose homeostasis.

We performed the ‘physical activity, myokines and glucose metabolism’ (MyoGlu) study ^29^, which was a controlled clinical trial aiming to identify novel secreted factors (‘exerkines’) that may serve as links between physical activity and glucose metabolism. We conducted a comprehensive serum screening of 3072 proteins in normal weight and overweight men both before and after combined endurance and strength exercise. Rigorous phenotyping was carried out, including hyperinsulinemic euglycemic clamping, assessments of maximum oxygen uptake, maximum muscle strength, and ankle-to-neck MRI/MRS scans.

Exerkines identified with potential effects on glucose homeostasis in the MyoGlu study were subsequently subject to analysis across several external data sets. Using data from 47,747 participants in the UK Biobank ^30^, we assessed correlations between candidate proteins and estimates of physical activity and glucometabolic outcomes. These associations were then tested for causality using Mendelian randomization (MR). Exerkines of interest were also assessed in a knockout mouse model and in a exercise mouse model to further assess potential links with glucose homeostasis.

## Results

### Cohort characteristics

We studied 26 male participants, including 13 with normal weight, and another 13 with overweight, as described previously ^29^. They were subjected to 12-weeks of high-intensity resistance and endurance exercise (Figure 1). The overweight participants had lower glucose tolerance and insulin sensitivity compared to the normal weight participants (Supplementary Table 1). After the 12-week intervention, body fat mass decreased and lean body mass increased, together with significant improvements in insulin sensitivity (∼40%), maximum oxygen uptake and muscle strength in both groups (Supplementary Table 1).

**Figure 1.**
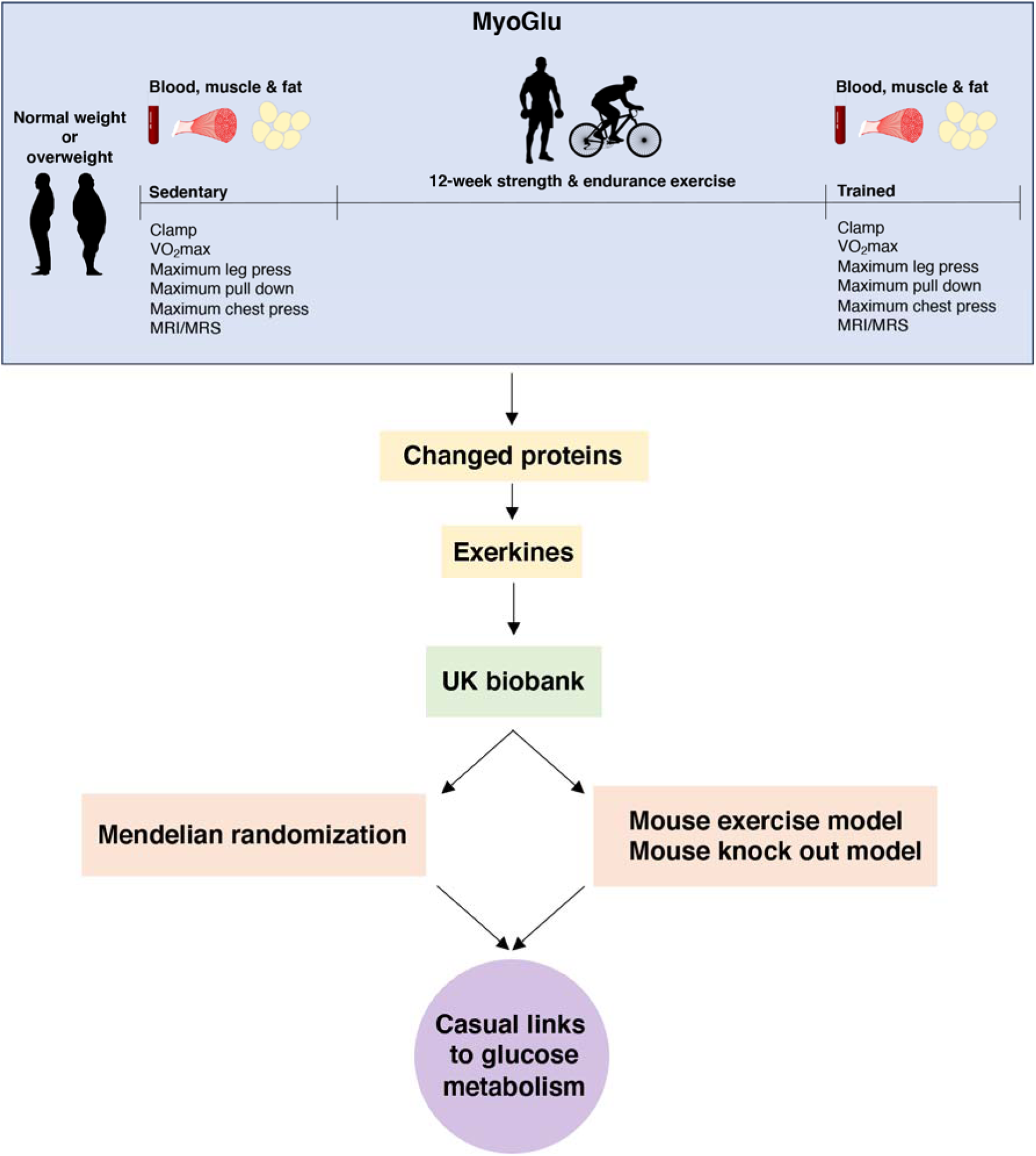
Study overview. We recruited sedentary men with either normal weight or overweight for deep phenotyping before and after a prolonged exercise intervention. Multiomic analyses, including serum proteomics, clinical traits and muscle and fat transcriptomics identified changed proteins and potential exerkines. Candidate exerkines were subsequently analyzed in serum samples from the UK biobank and tested for associations with physical activity and glucometabolic traits. Top candidates were then subjected to Mendelian randomization and investigated in a mouse exercise model and in a mouse knock-out model to assess casual links between exerkines and glucometabolic traits.

### Serum proteome responses to prolonged exercise

Recognizing that circulating proteins could mediate exercise-induced metabolic improvements, we next investigated alterations in the serum proteome in response to the 12-week intervention using PEA technology. Of the 3072 proteins quantified, we detected increased serum concentrations of 126 proteins, and decreased serum concentrations of 157 proteins following the 12-week intervention, at a false discovery rate (FDR) below 5% (Figure 2 A-C; Supplementary Table 2-4). Among these, 20 proteins increased exclusively in normal weight men, whereas 19 proteins increased exclusively in overweight men (Figure 2 D). Four proteins were uniquely reduced in normal weight men, and 66 proteins were uniquely reduced in overweight men (Figure 2 E).

**Figure 2.**
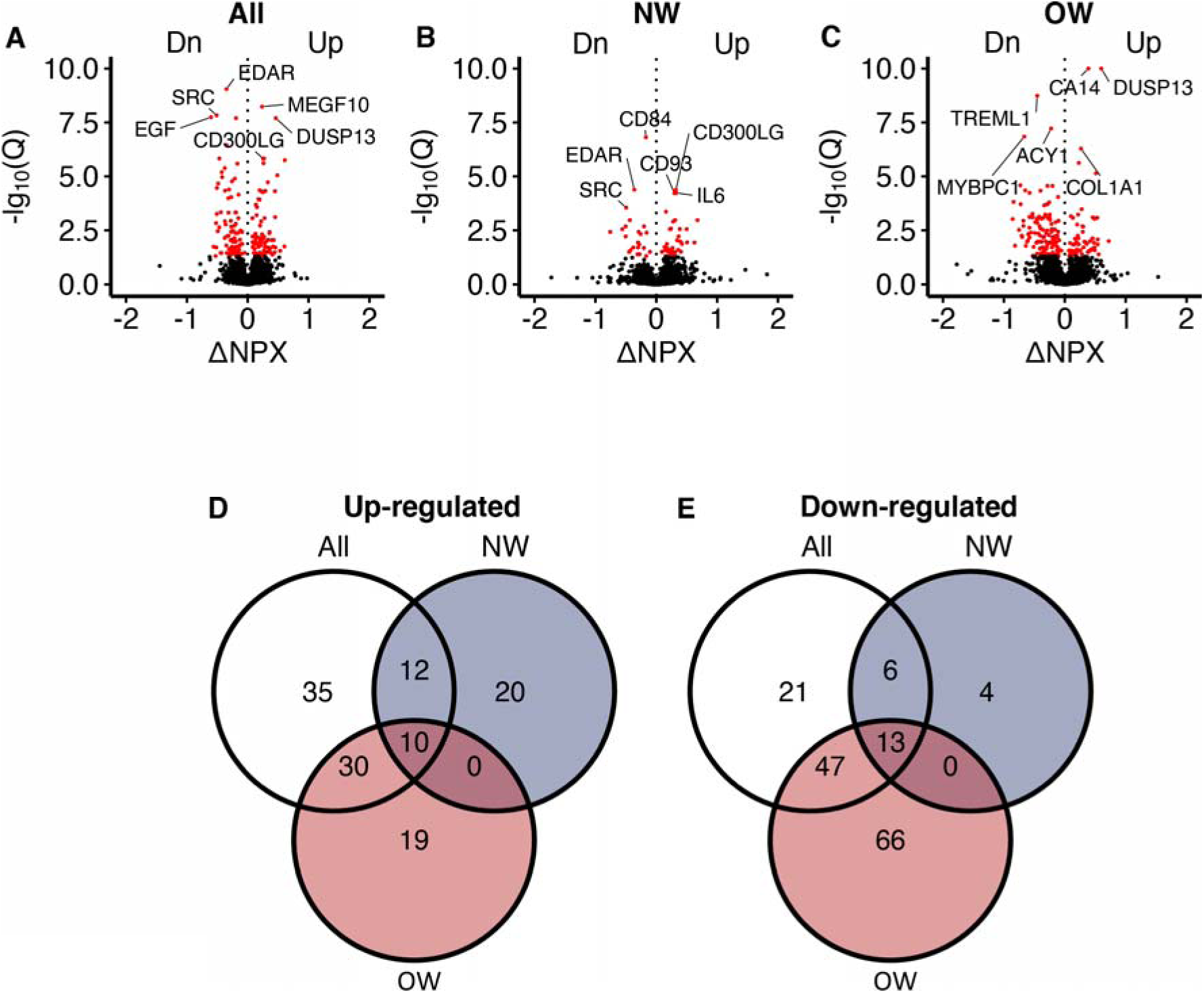
Serum proteomic responses to prolonged exercise. (A) A volcano plot showing responses in all participants. The X-axis shows log_2_(fold-changes) and the Y-axis shows negative log_10_(Q-values). The red dots indicate statistical significance (Q<0.05). Only the top 3 up/down-regulated proteins are annotated. (B-C) Similar to A, but in normal weight and overweight men only. (D-E) Venn diagrams of the significant change in proteins shown in A-C. NPX = normalized protein expression. Q = P-values corrected using Benjamini-Hochberg’s method. NW = normal weight. OW = overweight.

Several of the exercise-responsive proteins had potential roles in muscle adaptation and metabolism. For example, platelet-derived growth factor subunit B (PDGFB) and IL7, are both myokines with potential effects on muscle differentiation ^15,31^. Further, fibroblast growth factor-binding protein 3 (FGFBP3) may influence running capacity ^32^ and muscle strength ^33^. NADH-cytochrome b5 reductase 2 (CYB5R2) can preserve SkM mitochondria function in aging mice ^34^. FGFBP3 and switch-associated protein 70 (SWAP70) may protect against weight gain ^35^ and cardiac hypertrophy ^36^, respectively. Finally, dual specificity protein phosphatase 13 isoform A (DUSP13A) is highly specific to SkM ^37^, making it a potential novel muscle-specific marker for long-term exercise. Detailed results for 2885 proteins in response to prolonged exercise are shown in Supplementary Table 2.

### A proteomic liver fat signature in overweight men

In response to the 12-week exercise intervention, a larger number of serum proteins responded in overweight men than in normal weight men (Figure 2 B-C). In particular, 66 proteins decreased in serum after 12 weeks in overweight men (Figure 2 E). Gene ontology analyses revealed known pathways only for the proteins that decreased in overweight men (Figure 3 A-B), and one of the most enriched pathways is related to metabolism of organic acids (Figure 3 C). A key driver analysis of the 66 proteins identified SLC22A1, a hepatocyte transporter related to liver fat content (Figure 3 D). Furthermore, the 66 proteins also displayed a 24% overlap with a known human serum proteomic signature of non-alcoholic fatty liver disease (NAFLD; Figure 3 E) ^38^, but no common proteins with signatures of specific liver cells (The Human Liver Cell Atlas: ^39^). Baseline serum protein concentrations in the identified signature of 66 proteins were higher among men with overweight compared to those with normal weight, but were reduced or normalized in overweight men following prolonged exercise (Figure 3 F). Using principal component analysis of the 66 proteins, the first component correlated positively to liver fat content at baseline (Figure 3 G), but not after prolonged exercise (Figure 3 H). Similarly, the first component also correlated positively with several liver-related markers at baseline (Figure 3 L) and negatively to insulin sensitivity at baseline (Figure 3 I), but not after prolonged exercise (Figure 3 J). The first component mediated 37% of the association between baseline insulin sensitivity and liver fat content (Figure 3 K). We observed no enrichments for the remaining proteins (Figure 3 A-B).

**Figure 3.**
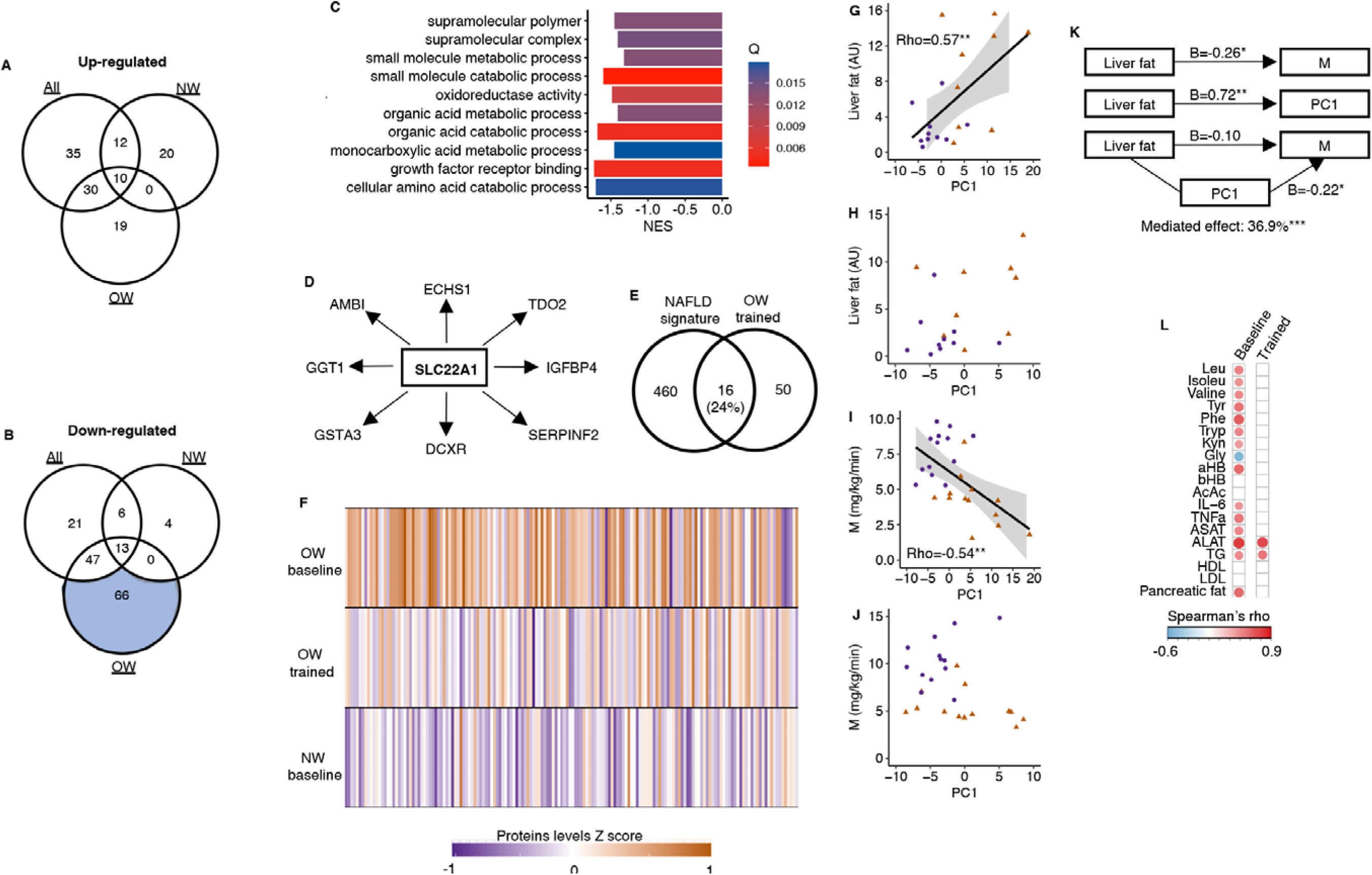
A serum proteomic liver fat signature. (A) No up-regulated proteins after prolonged exercise overlapped with known pathways. (B) Only the 66 down-regulated proteins in the OW group overlapped with known pathways. (C) Top 10 gene sets overlapping with these66 proteins. (D) SLC22A1 is a key driver among these 66 proteins. (E) These 66 proteins overlapped with a known human serum proteomic non-alcoholic fatty liver disease signature from Govaere et al^38^. (F) The down-regulated proteins in the OW group were elevated in OW vs. NW at baseline but normalized in the OW group after prolonged exercise. The principal component of these 66 proteins correlated with(G) liver fat content at baseline, but (H) not after prolonged exercise, with (I) the clamp M value at baseline, but (J) not after prolonged exercise. (K) The principal component (PC) of these 66 proteins mediated 36.9% of the association between liver fat and M. (L) The principal component of these 66 proteins correlated with several liver-related markers at baseline, but not after prolonged exercise except for ASAT and ALTA(white = non-significant, red/blue = significant). *p<0.05 and **p<0.01.

### Secretory proteins

Among the 96 up-regulated and 110 down-regulated serum proteins responding to the 12-week exercise intervention (Figure 2 D-E), 37 are curated secretory proteins, and another 46 proteins are predicted as highly likely secretory proteins (Figure 4 A-C). We assessed the corresponding mRNA responses in SkM and subcutaneous white adipose tissue (ScWAT) following the 12-week intervention (Figure 4 A-C). In total, 19.7 % of the serum secretory proteins displayed a directionally consistent significant change mRNA levels in SkM, whereas 12.1 % of the serum secretory proteins exhibited a corresponding mRNA response in ScWAT (Figure 4 C). *COL1A1* was the most responsive SkM mRNA that also had a corresponding increase in serum COL1A1 after prolonged exercise (Figure 4 D-E). *CCL3* was the most responsive ScWAT mRNA that also had a corresponding decrease in serum after prolonged exercise (Figure 4 F-G). To prioritize proteins for follow-up analyses, we focused on SMOC1 and CD300LG, which had similar exercise responses in blood, SkM and ScWAT (Figure 4 H). SMOC1 is a known hepatokine with effects on insulin sensitivity in mice ^40^, but probably with no causal link to insulin sensitivity in humans ^40,41^. Thus, we focused on CD300LG in subsequent analyses.

**Figure 4.**
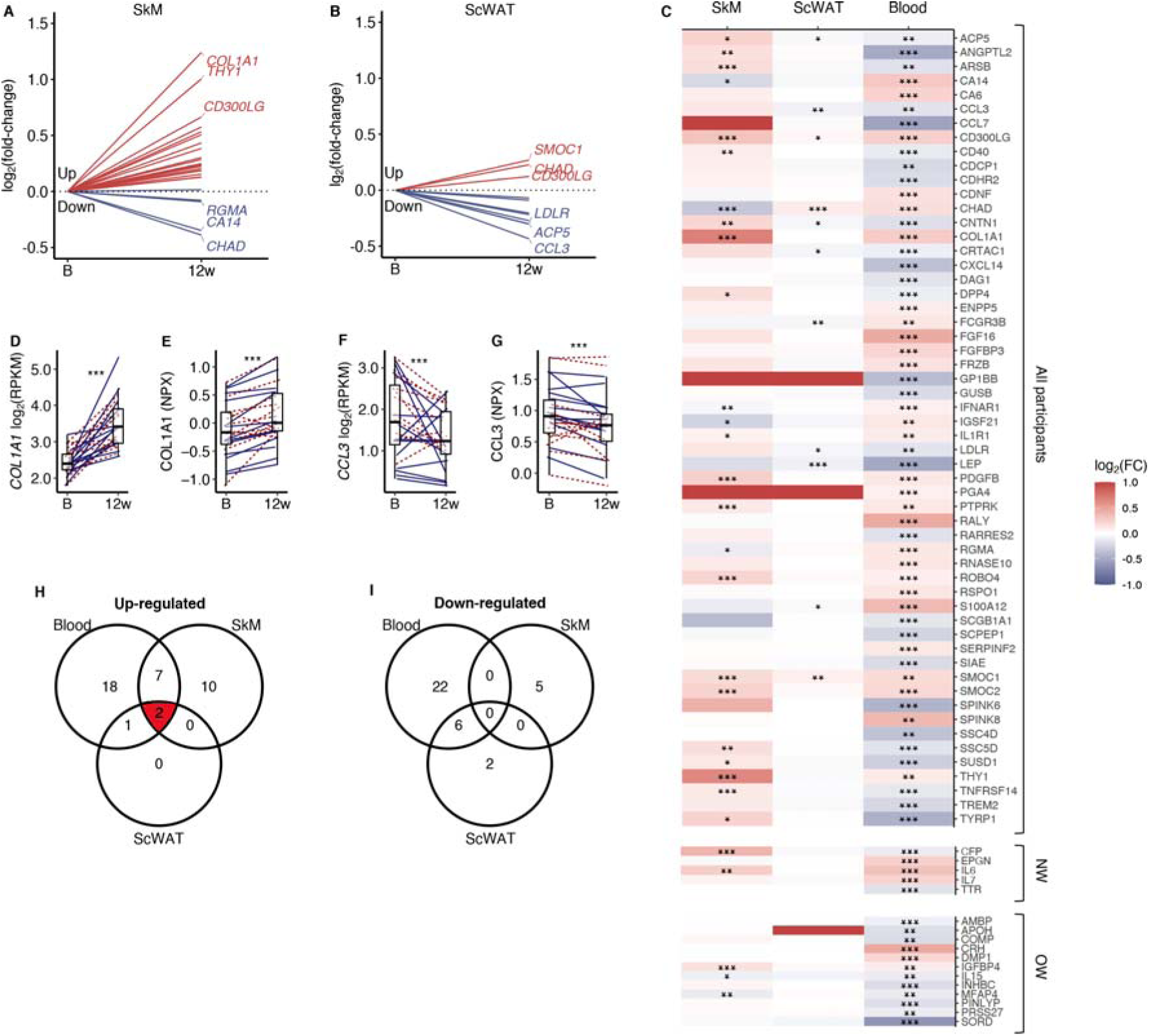
Comparison of secretory protein responses to prolonged exercise in blood with corresponding mRNA levels in skeletal muscle and adipose tissue. (A) mRNA levels in skeletal muscle and (B) adipose tissue for proteins that responded significantly to prolonged exercise. (C) A heatmap of log_2_(fold-changes) in blood, skeletal muscle and adipose tissue. (D) The most responding mRNA in skeletal muscle, and (E) the response in the blood protein. (F) The most responding mRNA in adipose tissue, and (G) the response in the blood. (H-I) Venn diagrams of significant changes in blood, skeletal muscle and adipose tissue. FC = fold-change. SkM = skeletal muscle. ScWAT = subcutaneous adipose tissue. NPX = normalized protein expression. RPKM = reads per kilobase per million mapped read. *p<0.05, **p<0.01 and ***p<0.001.

### CD300LG

CD300LG displayed increased concentration in serum (+20%, p<0.001) together with increased levels in both SkM (+60%, p<0.001) and scWAT (+13%, p=0.01) mRNA following the 12-week exercise intervention (Figure 5 A-C). Changes in serum CD300LG correlated positively with changes in insulin sensitivity after the intervention (rho=0.59, p=0.002; Figure 5 D). In addition, serum CD300LG concentration was lower in overweight than normal weight men (−51%, p=0.014) and positively correlated with insulin sensitivity before as well as after the 12-week intervention (pre-trained: r=0.50, p=0.001, and post-trained: r=0.43, p=0.028).

**Figure 5.**
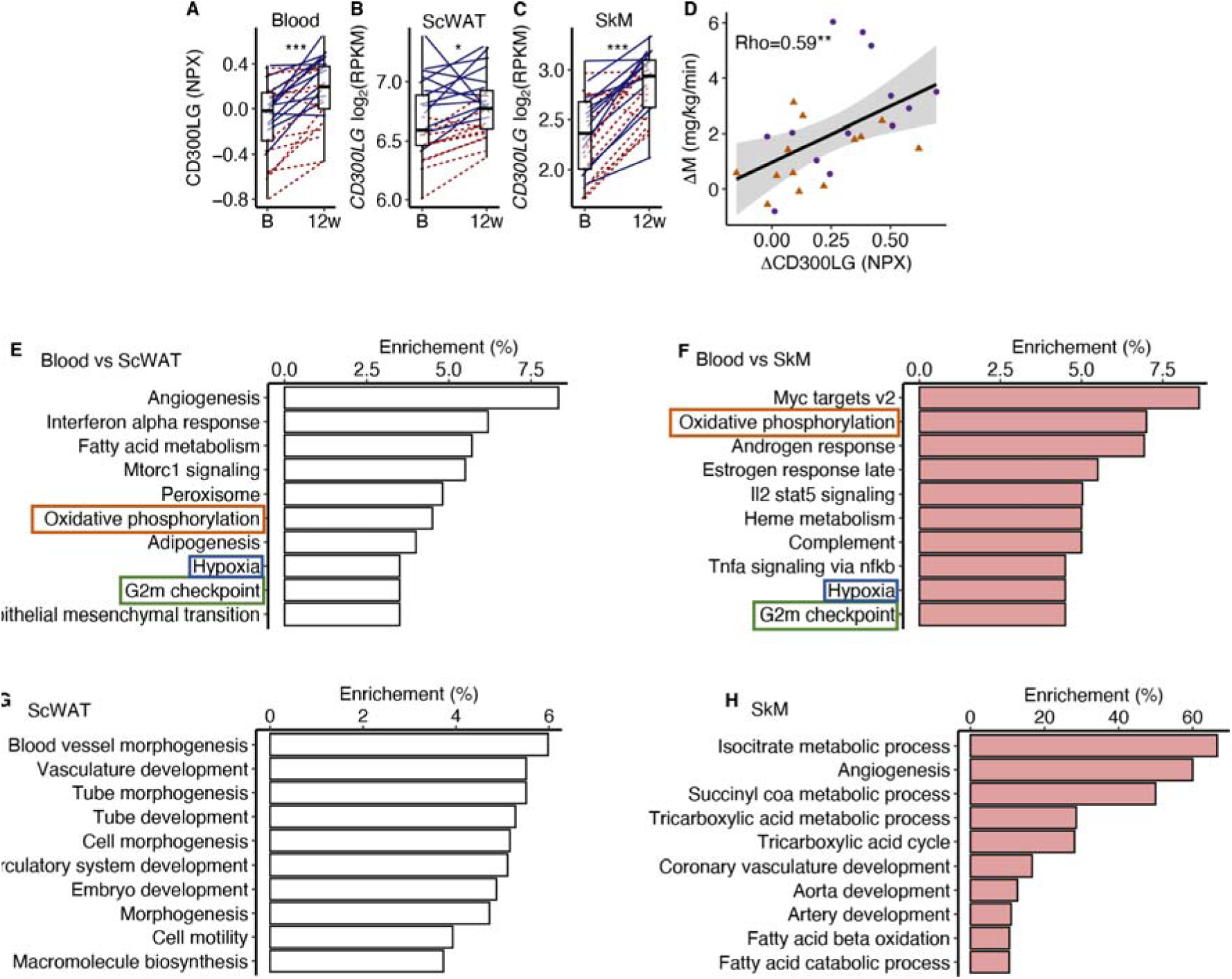
CD300LG. (A) The response from baseline to week 12 in serum CD300LG and CD300LG mRNA in (B) subcutaneous adipose tissue (ScWAT) and (C) skeletal muscle (SkM). (D) Correlation between the change from before to after prolonged exercise in serum CD300LG and insulin sensitivity. (E-H) Pathway enrichment analyses were performed on the top 500 most correlated (and p<0.05) genes in (E) ScWAT or (F) SkM to the change in serum CD300LG levels, or to the change in CD300LG mRNA levels in (G) ScWAT or (H) SkM. Only the top 10 pathways with Q<0.05 are presented. *p<0.05, **p<0.01 and ***p<0.001.

To investigate the potential effect of serum CD300LG on SkM and ScWAT, we performed an overrepresentation analysis on the top 500 mRNAs that were positively correlated (p<0.05) with serum CD300LG levels in each tissue (Figure 5 E-H). Pathway analyses revealed that the change in serum CD300LG concentrations correlated with changes in expression of genes involved in oxidative phosphorylation, G2M check point and hypoxia both in ScWAT and SkM (Figure 5 E-F). In ScWAT, serum CD300LG levels also showed the strongest enrichment with angiogenesis pathways (Figure 5 E). In ScWAT, the change in ScWAT *CD300LG* mRNA levels correlated positively with the change in ScWAT mRNA of genes related to angiogenesis/vasculature development (Figure 5 G). Similar correlations between *CD300LG* mRNA and angiogenesis genes were observed in SkM as well (Figure 5 H). For example, 60% of the mRNAs in the angiogenesis pathway correlated with *CD300LG* (Figure 5 H). However, serum CD300LG levels were also correlated positively with pathways related to fatty acid metabolism in both ScWAT (Figure 5 E) and SkM (Figure 5 H).

To explore tissue specific expression of CD300LG, we utilized data from a publicly available human tissue panel ^42^. CD300LG is highly expressed in adipose tissue compared to other tissues (Supplementary Figure 2 A) supporting our observation that ScWAT expression was substantially higher than in SkM (Figure 5 B-C). To further investigate which cells in ScWAT that express CD300LG, we utilized data from a single cell mRNA sequencing atlas of human ScWAT (https://singlecell.broadinstitute.org/single_cell) generated by Emont *et al.* ^43^. *CD300LG* mRNA in ScWAT was primarily expressed in venular endothelial cells, but not adipocytes or other cell types present in ScWAT (Supplementary Figure 2 B-E).

We next explored whether CD300LG mediates tissue-tissue cross-talk using data from the GD-CAT (Genetically-Derived Correlations Across Tissues) database ^44^ ^45^, which is a tool for analyzing human gene expression correlations in and across multiple tissues. In men, ScWAT *CD300LG* correlated strongly with ScWAT, SkM and aortic gene expression (Supplementary Figure 3). Consistent with our observations in the MyoGlu exercise intervention study, the top network of gene expression in ScWAT related to ScWAT *CD300LG* mRNA was angiogenesis (Supplementary Figure 3 A). Like ScWAT, SkM *CD300LG* also correlated strongly with ScWAT, SkM and aortic gene expression (Supplementary Figure 3 B). The proteasome complex was the top network of gene expression related to SkM *CD300LG* mRNA Supplementary Figure 3 B). In contrast, running the same analyses in women did not reveal associations between *CD300LG* and angiogenesis (Supplementary Figure 4 A-B).

We then evaluated serum CD300LG levels in up to 47,747 samples in the UK biobank (see Methods). Descriptive statistics of the UK biobank cohort are presented in Supplementary Table 5. Serum CD300LG levels were positively associated with several measures of physical activity (all metabolic equivalent tasks, results from the international physical activity questionnaire and meeting the recommended amount of weekly physical activity or not; Table 1). Interestingly, serum CD300LG levels were most strongly related to vigorous activity (Table 1). Furthermore, the associations between serum CD300LG and physical activity were significantly stronger in men than in women (Table 1). Serum CD300LG levels were also positively associated with fat mass and fat free mass, and negatively associated with glucometabolic traits including serum glucose levels, Hb1Ac and the risk of having type 2 diabetes (Table 1). These associations were independent of body mass index (BMI).

**Table 1.** Multiple regression analyses between serum CD300LG, and measures of physical activity and glucometabolic traits in the UK biobank.

|  | Women |  |  | Men |  |  | Interaction |  |  | Description | No. of women | No. of men |
| --- | --- | --- | --- | --- | --- | --- | --- | --- | --- | --- | --- | --- |
|  | Beta-estimate | SE | P | Beta-estimate | SE | P | Beta-estimate | SE | P |  |  |  |
| NPX ~ physical activity |  |  |  |  |  |  |  |  |  |  |  |  |
| MET per week all activity | 1,0E-06 | 1,2E-06 | 0,384 | 4,5E-06 | 9,9E-07 | <0.001 | 4,3E-06 | 1,5E-06 | 0,005 | MET minutes per week | 22527 | 20726 |
| MET minutes walking | -6,0E-07 | 2,6E-06 | 0,818 | -1,8E-06 | 2,6E-06 | 0,478 | 1,7E-07 | 3,7E-06 | 0,962 | MET minutes per week | 22527 | 20726 |
| MET minutes moderate activity | -2,5E-06 | 2,4E-06 | 0,294 | 1,9E-07 | 2,3E-06 | 0,932 | 1,9E-06 | 3,3E-06 | 0,554 | MET minutes per week | 22527 | 20726 |
| MET minutes vigorous activity | 1,0E-05 | 2,8E-06 | <0.001 | 2,2E-05 | 2,1E-06 | <2e-16 | 1,5E-05 | 3,5E-06 | <0.001 | MET minutes per week | 22527 | 20726 |
| Sedentary overall average | 0,077 | 0,053 | 0,147 | 0,042 | 0,054 | 0,441 | -0,100 | 0,075 | 0,185 | Proportion sedentary activity. | 7430 | 5825 |
| Light overall average | -0,119 | 0,062 | 0,053 | -0,420 | 0,071 | <0.001 | -0,232 | 0,094 | 0,013 | Proportion light activity. | 7430 | 5825 |
| Moderate/vigorous overall average | 0,633 | 0,171 | <0.001 | 1,357 | 0,194 | <0.001 | 1,043 | 0,254 | <0.001 | Proportion moderate/vigorous activity. | 7430 | 5825 |
| IPAQ activity group | 9,6E-03 | 3,8E-03 | 0,012 | 3,1E-02 | 3,8E-03 | <0.001 | 2,6E-02 | 5,4E-03 | <0.001 | IPAQ category | 22527 | 20726 |
| Summed days activity | 1,9E-03 | 5,9E-04 | <0.001 | 3,8E-03 | 5,7E-04 | <0.001 | 2,7E-03 | 8,1E-04 | <0.001 | Days performing walking, moderate and vigorous activity | 23199 | 21138 |
| Summed minutes activity | -1,8E-05 | 3,0E-05 | 0,550 | 6,4E-05 | 2,7E-05 | 0,017 | 9,9E-05 | 4,0E-05 | 0,013 | Mins performing walking, moderate and vigorous activity | 22527 | 20726 |
| Moderate/vigorous recommendation1 | 7,5E-03 | 5,6E-03 | 0,186 | 4,8E-02 | 5,8E-03 | < 2e-16 | 4,6E-02 | 8,0E-03 | <0.001 | Yes/No | 22527 | 20726 |
| Moderate/vigorous walking recommendation1 | -1,3E-05 | 7,2E-03 | 0,999 | 4,0E-02 | 7,4E-03 | <0.001 | 4,6E-02 | 1,0E-02 | <0.001 | Yes/No | 22521 | 20723 |
| Trait ~ NPX |  |  |  |  |  |  |  |  |  |  | 28108 | 23841 |
| Body fat percentage impedance | -0,137 | 0,051 | 0,007 | -0,552 | 0,052 | <0.001 | -0,375 | 0,073 | <0.001 | Body fat percentage | 28099 | 23802 |
| Whole body fat mass impedance | 0,345 | 0,049 | <0.001 | 0,023 | 0,051 | 0,656 | -0,209 | 0,071 | 0,003 | Fat mass (kg) | 28108 | 23866 |
| Whole body fat free mass impedance | 0,450 | 0,050 | <0.001 | 1,045 | 0,090 | <0.001 | 0,622 | 0,101 | <0.001 | Fat free mass (kg) | 28107 | 23870 |
| Body mass index | - | - | - | - | - | - | - | - | - | kg/m2 | 24810 | 21509 |
| Glucose | -0,066 | 0,016 | <0.001 | -0,041 | 0,022 | 0,062 | 0,033 | 0,026 | 0,202 | mmol/L | 27271 | 23294 |
| HbA1c | -0,898 | 0,078 | <0.001 | -0,911 | 0,107 | <0.001 | -0,010 | 0,130 | 0,936 | mmol/mol | 27323 | 23292 |
| Triglycerides | -0,399 | 0,011 | <0.001 | -0,413 | 0,017 | <0.001 | -0,058 | 0,020 | 0,004 | mmol/L | 28387 | 24332 |
| Type 2 diabetes | -0,012 | 0,002 | <0.001 | -0,018 | 0,004 | <0.001 | 0,000 | 0,004 | 0,982 | Yes/No. | 24802 | 21483 |
| TyG | -2,189 | 0,074 | <0.001 | -2,228 | 0,120 | <0.001 | -0,203 | 0,136 | 0,138 | mmol/L x mmol/L | 22527 | 23841 |

MET = metabolic equivalent of task. NPX = Normalized protein expression. SE = standard error. IPAQ = International physical activity questionnaire. TyG = triglycerid glucose index on insulin resistance.
Model 1 (NPX – physical activity) was a linear regression model of NPX values as a function of a measure of physical activity.
Model 2 (Trait – NPX) the measures of body composition and glucometabolic traits were the outcomes and NPX values were set as the exposure.
Models 1 and 2 were adjusted for age, batch, study centre, storage time and BMI.
<sup>1</sup>Indicates whether a person met the 2017 UK Physical activity guidelines of 150 minutes of moderate activity per week or 75 minutes of vigorous activity.

### GWAS of serum CD300LG levels

GWAS analyses of CD300LG levels detected 43 independent genome-wide significant genetic associations across the genome (Supplementary Figure 5). The genomic inflation factor (λ=1.0966) and LD score intercept (1.039) were consistent with our GWAS being well controlled for population stratification and other possible biases (Supplementary Figure 6). The most significant SNPs lay along chromosome 17 with these SNPs mapping to the genomic region encoding the *CD300LG* gene (Supplementary Figure 5). Follow-up analyses revealed three significant, independent *cis*-pQTLs associated with the protein CD300LG (Supplementary Table 6) and a number of *trans*-pQTLs (Supplementary Table 7).

#### Mendelian randomization (MR) analysis

The independent genome-wide significant SNPs from the CD300LG GWAS were used for two-sample MR (see Methods for details), where 39 SNPs were also available in the outcome GWAS ^46^. We first performed Inverse Variance Weighted (IVW) MR analysis to test the causal relationship between CD300LG and fasting glucose, 2-hour post oral glucose tolerance test (OGTT) glucose levels and HbA1c using only *cis*-SNPs (Supplementary Table 8) and all SNPs (Supplementary Table 9). The *cis* IVW MR analysis showed some evidence for a negative causal effect of CD300LG on fasting insulin (p=0.01), but due to only three SNPs in these analyses, we could not perform additional sensitivity analyses (except for tests of heterogeneity in estimates of the causal effect across SNPs) and could not determine whether the absence of strong evidence for a causal effect on the glycaemic parameters was genuine or whether our analyses just lacked power. Although some of the analyses involving all the genome-wide significant SNPs indicated a potential causal link between increased serum CD300LG concentration and these outcomes (Supplementary Table 9) the analysis showed significant heterogeneity. We did not detect strong evidence of directional pleiotropy (significant MR Egger intercept, Supplementary Table 9). The heterogeneity in the analysis is possibly due to the fact that many of the SNPs found in the GWAS of CD300LG are associated with related phenotypes which could exert pleiotropic effects on diabetes related outcomes, and so the results should be interpreted with care (Supplementary Table 10). Due to the heterogeneity in our results we therefore performed MR PRESSO to account for outliers. The MR PRESSO analysis showed a significant negative effect of CD300LG on all outcomes (Table 2).

**Table 2.** Mendelian randomization of serum CD300LG levels and glucose outcomes using MR PRESSO.

| Outcome | MR analysis | Number of outliers | Effect | SD | p-value |
| --- | --- | --- | --- | --- | --- |
| 2-hour post OGTT glucose (mmol/L) | Raw | | -0.3722 | 0.0998 | $6.2 \times 10^{-4}$ |
| 2-hour post OGTT glucose (mmol/L) | Outlier-corrected | 2 | -0.3049 | 0.0855 | $1.04 \times 10^{-2}$ |
| Fasting glucose (mmol/L) | Raw |  | -0.0307 | 0.0358 | 0.3963 |
| Fasting glucose (mmol/L) | Outlier-corrected | 2 | -0.0556 | 0.0133 | $1.73 \times 10^{-4}$ |
| Fasting insulin (pmol/L) | Raw |  | -0.0870 | 0.0558 | 0.1271 |
| Fasting insulin (pmol/L) | Outlier-corrected | 10 | -0.0534 | 0.0252 | 0.0432 |
| HbA1c (%) | Raw | | -0.0485 | 0.0155 | $3.28 \times 10^{-3}$ |
| HbA1c (%) | Outlier-corrected | 3 | -0.0560 | 0.0155 | $1.04 \times 10^{-4}$ |

#### Mouse models

To functionally validate association of CD300LG with metabolic homeostasis, we leveraged phenotypic data for exercising mice and for *Cd300lg* deficient (*Cd300lg*^-/-^) mice that both were publicly available through the MoTrPAC ^47^ study and the international mouse phenotyping consortium (PhenoMouse) ^48^.

There is a 51% homology between human *CD300LG* and mouse *Cd300lg* ^49^, and also in mice, Cd300lg is predominantly expressed in adipose tissue endothelial cells ^43^.

In MoTrPAC ^47^, Cd300lg levels in scWAT from n=12-15 male and female mice were increased after exercise for 8 weeks (∼30% in both female (p=0.03) and male (p=0.01) mice) (Supplementary Figure 7 A-C). Based on data from n=3050 mice from PhenoMouse male, but not female, mutants for the *Cd300lg*^tm1a(KOMP)Wtsi^ allele displayed impaired glucose tolerance (Supplementary Figure 7 D), but no change in fasting glucose and insulin (Supplementary Figure 7 E-F). Mutant male, but not female, mice also displayed increased lean mass (Supplementary Figure 7 G) and less fat mass (Supplementary Figure 7 H). Detailed PhenoMouse results are presented in Supplementary Table 11.

## Discussion

In the present study, we characterized the effects of strength and endurance exercise on the serum proteome of sedentary normal weight and overweight men. We identified significant changes in 283 serum proteins related to many signaling pathways after the 12-week intervention. Some of these proteins were related to the mitochondria, muscle differentiation and exercise capacity. Among known secretory proteins, 19.7% and 12.1% displayed corresponding mRNA changes in SkM and ScWAT, respectively. Although some proteins may be myokines, others may be adipokines or other types of exerkines. A multi-tissue responding protein was CD300LG, which also correlated positively to insulin sensitivity. CD300LG was particularly interesting because we could replicate the finding in an external cohort, find evidence of a causal link to glucose homeostasis, and perform functionally validation in mice models. Furthermore, the association between CD300LG, physical activity and glycemic traits might display sex dimorphic relationships.

One of the protein signatures observed in response to exercise was based on strong associations with markers of liver fat content in overweight men. This was related to SLC22A1, which regulates the hepatic glucose-fatty acid cycle affecting gluconeogenesis and lipid metabolism ^50^, and may influence liver fat accumulation ^51^. This signature also shared many common proteins with a known serum NAFLD proteomic signature ^38^. However, we did not detect overlaps between proteins in this signature and specific gene expression patterns of liver cells (e.g. hepatocytes, immune cells etc.) ^39^. This observation suggests that the proteins detected do not relate to liver protein synthesis per se, but may accumulate in serum due to being released in the blood stream as a result of impaired liver protein catabolism or cell damage as a consequence of overweight/obesity. Notably, this protein signature in overweight men normalized after 12 weeks of exercise and resembled the signature observed in normal weight men. These data suggest prolonged exercise leads to improvements of liver function in overweight men.

Several proteins responding to prolonged exercise had a known signal sequence. These secretory proteins are of particular interest because they could mediate inter-tissue adaptations to exercise. For example, COL1A1 was substantially increased in serum and its corresponding mRNA level was increased in SkM. However, COL1A1 is a collagen peptide that is related to muscle damage, turn-over and extracellular matrix remodeling in response to exercise ^52^ and may mostly reflect muscle restructuring and not represent signaling effects to distant tissues. The large overlap between serum proteins and SkM mRNA most likely suggests a similar phenomenon, where tissue restructuring following exercise is reflected in blood. However, there are probably also several myokines with distant signaling effects among the identified proteins. CCL3 was reduced in serum in parallel with a reduction in its mRNA level in ScWAT. CCL3 is a monocyte chemoattractant protein that may be related to immune cell infiltration in adipose tissue ^53^. Hence, this may reflect a positive effect of prolonged exercise on adipose tissue inflammation, which is in line with our previous results showing normalization of adipose tissue inflammation following prolonged exercise ^10^.

A particularly interesting protein was CD300LG, which responded to prolonged exercise in serum, and, judged by its mRNA levels, in SkM and ScWAT. Serum CD300LG levels were lower in overweight compared to normal weight men. Furthermore, the exercise-induced response in CD300LG correlated positively to improvements in insulin sensitivity, and there was also a significant correlation between serum CD300LG and insulin sensitivity both before and after the intervention. We therefore analyzed CD300LG in an external data set, the UK Biobank, and again we observed positive associations between especially vigorous exercise and serum CD300LG. Moreover, serum CD300LG levels were negatively associated with glucose levels and type 2 diabetes in the UK Biobank, and these associations might be causal based on MR analysis. These findings were functionally corroborated by the alterations in glucose tolerance and parameters related to insulin sensitivity observed in *Cd300lg*^-/-^ mice. Thus, CD300LG may represent an exerkine with a causal link to glucose homeostasis. However, whether CD300LG can mediate tissue-tissue crosstalk is unknown. CD300LG is a cell surface protein with a transmembrane domain, but is also a predicted secretory protein ^54^. Whether the protein is released from the cell surface in a regulated manner to mediate cross-tissue signaling needs further investigation. Furthermore, the exact link between CD300LG and glucose metabolism is not clear, but possibly related to the fact that CD300LG is expressed in endothelial cells ^55^, linked to blood pressure ^56^, lymphocyte binding ^57^, blood triacylglycerol levels ^58,59^ and molecular traffic across the capillary endothelium ^60^. Both MyoGlu and GD-CAT data also suggested that CD300LG may be related to angiogenesis in ScWAT ^61^ and SkM ^61,62^, at least in men. Hence, we speculate that the link between CD300LG and glucose metabolism is related to improved tissue capillarization/vascular function following prolonged exercise. Furthermore, since vigorous exercise leads to angiogenesis in ScWAT and SkM ^61^, serum CD300LG may be a maker of exercise intensity.

### Strengths and limitations

Although MyoGlu included only 26 sedentary men, they were extensively phenotyped with euglycemic hyperinsulinemic clamp, fitness tests, whole body imaging (MRI/MRS) and mRNA sequencing of ScWAT and SkM. We also supplied our study with data from 47747 persons in the UK biobank to enhance the validity and generalization of the results. Furthermore, to assess sex differences we stratified analyses for men and women in the UK Biobank, in external data bases (GD-CAT^45^) and analyzed data from both male and female mice. Since correlations with the clamp data only imply a role for a protein with regard to glucose homeostasis, so we also tested associations with related glucometabolic traits in the UK Biobank and tested these associations for causality using MR. We also included data from exercised mice, and mutant mice to further strengthen the results. Our serum proteome study assessed 3072 proteins, and therefore we do not cover the complete human proteome. However, the Olink platform is based on dual recognition of correctly matched DNA-labeled antibodies, and DNA sequence-specific protein-to-DNA conversion to generate a signal. This is a highly scalable method with an exceptional specificity (https://olink.com/application/pea/). Previous exercise-proteomic studies has looked at ∼600 proteins in overweight men after endurance exercise ^25^, and three papers were published from the HERITAGE study analyzing ∼5000 proteins in response to endurance exercise ^26–28^. However, our study is the first and largest exercise study using PEA in both overweight and normal weight men, and also including strength exercise.

However, CD300LG’s role related to angiogenesis is only suggested through association analyses in our data, necessitating follow-up studies to confirm any causal role of CD300LG in angiogenesis. Although the open-source cd300lgtm1a(KOMP)Wtsi mice provided interesting indications, a future study should directly phenotype mice with alterations in the CD300LG gene, and measure the effects on circulating CD300LG levels and potential regulatory mechanisms related to angiogenesis and glucose tolerance. Furthermore, it is also unknown if circulating CD300LG is full-length or a cleaved fragment, and the mechanisms for CD300LG secretion should be further studied *in vitro*. Finally, future experiments should also identify the epitope for O-link binding, and confirm its specificity using targeted mass spectrometry or antibody-based validations.

### Conclusion

Our study provided a detailed analysis of serum proteins responding to three months of strength and endurance exercise in both normal weight and overweight men. Our results identified a novel NAFLD-related serum protein signature in overweight men that was normalized after prolonged exercise. We also identified hundreds of tissue-specific and multi-tissue serum markers of e.g., mitochondrial function, muscle differentiation, exercise capacity and insulin sensitivity. Our results were enriched for secretory proteins (exerkines), such as CD300LG, which may be a marker of exercise intensity especially in men, and may also have causal roles in improved glucose homeostasis after physical activity.

## Methods

The MyoGlu study was conducted as a controlled clinical trial (clinicaltrials.gov: NCT01803568) and was carried out in adherence to the principles of the Declaration of Helsinki. The study received ethical approval from the National Regional Committee for Medical and Health Research Ethics North in Tromsø, Norway, with the reference number 2011/882. All participants provided written informed consent before undergoing any procedures related to the study. The UK biobank has ethical approval from the North West Multi-Centre Research Ethics Committee (MREC), which covers the UK, and all participants provided written informed consent. This particular project from the UK biobank received ethical approval from the Institutional Human Research Ethics committee, University of Queensland (approval number 2019002705).

### Participants

The MyoGlu study enrolled men aged 40 to 65 years who were healthy but sedentary (having engaged in fewer than one exercise session per week in the previous year) ^29,63^. These participants were divided into two groups based on their BMI and glucose tolerance: overweight (with an average BMI of 29.5 ± 2.3 kg/m^2^) and normal weight controls (with an average BMI of 23.6 ± 2.0 kg/m^2^). The overweight men had reduced glucose tolerance and/or insulin sensitivity (Supplementary Table 1). Both groups, consisting of 13 individuals each, underwent a 12-week regimen of combined strength and endurance training.

### Exercise protocols

This 12-week training intervention included two weekly sessions of 60 minutes each for endurance cycling and two sessions of 60 minutes each for whole-body strength training. Prior to and after the 12-week exercise intervention, a 45-minute bicycle test at 70% of their maximum oxygen uptake (VO_2_max) was conducted. Serum, muscle (*m. vastus lateralis*) and subcutaneous white adipose tissue biopsies were taken before, and 48 hours after the last exercise session of the 12-week intervention period ^29,63^.

### Clinical data

The euglycaemic hyperinsulinemic clamp was performed after an overnight fast ^29,63^. A fixed dose of insulin 40 mU/m^2^·min^−1^ was infused, and glucose (200 mg/mL) was infused to maintain euglycaemia (5.0 mmol/L) for 150 min. Insulin sensitivity is reported as glucose infusion rate (GIR) during the last 30 min relative to body weight. Whole blood glucose concentration was measured using a glucose oxidase method (YSI 2300, Yellow Springs, OH) and plasma glucose concentration was calculated as whole blood glucose x 1.119. Magnetic resonance imaging/spectroscopy (MRI/MRS) methods were used to quantify fat and lean mass. The ankle-to-neck MRI protocol included a 3D DIXON acquisition providing water and lipid quantification, data were then analysed using the nordicICE software package (NordicNeuroLab, Bergen, Norway), and the jMRUI workflow. VO_2_max tests were performed after standardized warm-up at a workload similar to the final load of an incremental test in which the relationship between workload (watt) and oxygen uptake was established. Participants cycled for onelllmin followed by a 15 watt increased workload every 30llls until exhaustion. Test success was based on O_2_ consumption increased <0.5lllmL·kg^−1^·min^−1^ over a 30 watt increase in workload, respiratory exchange ratio values >1.10, and blood lactatelll>7.0lllmmol/L. We obtained scWAT, SkM biopsies and blood samples as described previously ^29^. Biopsies were obtained from the periumbilical subcutaneous tissue and from *m. vastus lateralis*. After sterilization, a lidocaine based local anaesthetic was injected in the skin and sub cutis prior to both SkM and scWAT biopsies. Biopsies were dissected on a cold aluminium plate to remove blood etc. before freezing. For standard serum parameters, measurement were either conducted using standard in-house methods or outsourced to a commercial laboratory (Fürst Laboratories, Oslo, Norway).

### The Olink proteomics explorer 3072 platform

We utilized antibody-based technology (Olink Proteomics AB, Uppsala, Sweden) to conduct profiling of serum samples through the Olink Explore 3072 panel. This PEA technique involves using pairs of DNA oligonucleotide-labeled antibodies to bind to the proteins of interest. When two matching antibodies attach to a target protein, the linked oligonucleotides hybridize and are extended by DNA polymerase, forming a unique DNA “barcode”. This barcode is then read using next-generation sequencing. The specificity and sensitivity of the PEA technology are notably high because only accurately matched DNA pairs generated detectable and measurable signals. To refine the dataset, proteins that were not detected or were duplicated were removed, resulting in an analysis of 2886 proteins. Only a single assay was conducted, eliminating inter-assay variability. Data are presented as normalized protein expression (NPX) units, which are logarithmically scaled using a log_2_ transformation.

### Proteomics validations

Duplicate measurements of IL6 and leptin in plasma were conducted using ELISA kits (Leptin; Camarillo, CA and IL6; R&D Systems, Minneapolis, MN) following the manufacturer’s instructions. The correlations between PEA or enzyme linked immunosorbent (ELISA) assays were r=0.94 (p=1.4×10^−11^), and r=0.92 (p=2.2×10^−11^) for IL6 and leptin, respectively (Supplementary Figure 1).

### mRNA sequencing

Biopsies were frozen in liquid nitrogen, crushed to powder by a pestle in a liquid nitrogen-cooled mortar, transferred into 1 mL QIAzol Lysis Reagent (Qiagen, Hilden, Germany), and homogenized using TissueRuptor (Qiagen) at full speed for 15 sec, twice ^29,63^. Total RNA was isolated from the homogenate using miRNeasy Mini Kit (Qiagen). RNA integrity and concentration were determined using Agilent RNA 6000 Nano Chips on a Bioanalyzer 2100 (Agilent Technologies Inc., Santa Clara, CA). RNA was converted to cDNA using High-Capacity cDNA Reverse Transcription Kit (Applied Biosystems, Foster, CA). The cDNA reaction mixture was diluted in water and cDNA equivalent of 25 ng RNA used for each sample. All muscle and scWAT samples were deep-sequenced using the Illumina HiSeq 2000 system with multiplex at the Norwegian Sequencing Centre, University of Oslo. Illumina HiSeq RTA (real-time analysis) v1.17.21.3 was used. Reads passing Illumina’s recommended parameters were demultiplexed using CASAVA v1.8.2. For prealignment quality checks, we used the software FastQC v0.10.1. The mean library size was ∼44 millions unstranded 51 bp single-ended reads for muscle tissue and ∼52 millions for scWAT with no differences between groups or time points. No batch effects were present. cDNA sequenced reads alignment was done using Tophat v2.0.8, Samtools v0.1.18, and Bowtie v2.1.0 with default settings against the UCSC hg19 annotated transcriptome and genome dated 14^th^ of May 2013. Post-alignment quality controls were performed using the Integrative Genome Viewer v2.3 and BED tools v2.19.1. Reads were counted using the intersection strict mode in HTSeq v0.6.1.

### Statistics and bioinformatics

Olink data were analyzed using the ‘AnalyzeOlink’ R package for pre-processing, testing using mixed linear regression and annotation. Pathway and gene ontology overrepresentation analyses were performed using MSigDB data sets (Hallmark pathways and biological processes). mRNA sequencing data were normalized as reads per kilobase per million mapped read (RPKM) and analyzed using mixed linear regression from the ‘lme4’ R package. Normality was determined by quantile-quantile plots. P-values were corrected using the Benjamini-Hochberg approach set at a false discovery rate (FDR) of 5%. For univariate correlations, Pearsons’ or Spearmans’ method were applied as appropriate. Principal component analysis was performed using the ‘prcomp’ R package. Key driver analysis was performed using the ‘Mergeomics’ R package. Mediation analysis was performed using the ‘Mediation’ R package with 1000 bootstraps and the *set.seed* function to ensure reproducibility.

### UK Biobank

The UK biobank is a large prospective population-based cohort containing ∼500,000 individuals (∼273,000 women), with a variety of phenotypic and genome-wide genetic data available ^64^. The UK biobank has ethical approval from the North West Multi-Centre Research Ethics Committee (MREC), which covers the UK, and all participants provided written informed consent.

We utilized imputed genetic data from the October 2019 (version 3) release of the UK biobank for our analyses (Application ID: 53641). In addition to the quality control metrics performed centrally by the UK biobank^65^, we defined a subset of unrelated “white European” individuals. We excluded those with putative sex chromosome aneuploidy, high heterozygosity or missing rate, or a mismatch between submitted and inferred sex as identified by the UK biobank (total N = 1932). We excluded individuals who we did not identify as ancestrally European using K-means clustering applied to the first four genetic principal components generated from the 1000 Genomes Project ^66^. We also excluded individuals who had withdrawn their consent to participate in the study as of February 2021.

### The Olink proteomics explorer 1536 platform

All analysis were done using the UK Biobank Olink data containing a total of 58699 samples and 54309 individuals, after excluding individuals as mentioned above we had 47747 samples with measured serum CD300LG levels. Data was generated according to Olink’s standard procedures.

### Observational analyses

For the physical activity measurements we investigated if the degree of physical activity was associated with serum levels of protein (serum levels of protein regressed on physical activity), alternatively for the metabolic measurements we investigated if the protein expression affected the metabolic measurements (trait regressed on serum levels of protein), for both we used a linear regression model. We performed analyses stratified by sex and adjusting for age, protein batch, UK Biobank assessment centre, the time the sample was stored and BMI. All analyses were performed in R version 3.4.3.

### Genome-wide association analysis

A GWAS of serum CD300LG levels (log_2_ transformed) measured in the UK Biobank was performed using BOLT-LMM ^67^ on individuals of European descent who had proteomic data available (N=45788). We included sex, year of birth, protein and genotyping batch, time from sample collection to processing time (in weeks) and five ancestry informative principal components as covariates in the analysis.

Post GWAS quality control included the removal of SNPs with MAF ≤0.05 and info score ≤0.4 (n_SNPs_=6,945,819). The previously generated LD reference panel for clumping consisted of a random sample of 47674 unrelated British UK biobank individuals identified using GCTA ^68^ with identity by state (IBS)<0.025 and identity by descent (IBD) sharing oflll<0.1. LD score regression analysis ^69^ was used to investigate whether genomic inflation was likely due to polygenicity or population stratification/cryptic relatedness.

Prior to gene annotation palindromic SNPs were excluded (n_SNPs_=6,882,889 remaining). Variants were classified as either *cis-* or *trans-*pQTLs based on SNP proximity to the protein-encoding gene (CD300LG) of interest. Variant annotation was performed using ANNOVAR ^70^, labelling genes +/-500kb from variants. A pQTL was considered a cis-pQTL if the gene annotation in the 1Mb window matched the protein name, all remaining variants were considered trans-pQTLs.

To extract independent genome-wide significant pQTLs (*p*<5lll×lll10^−8^) clumping was performed using the PLINK v1.90b3.31 software package ^71^; variants with r^2^>0.001 with the index SNP were removed using a 1 Mb window. Variants which lied within the human major histocompatibility complex (MHC) region were removed, excluding pQTLs on chromosome 6 from 26Mb to 34Mb.

### Mendelian Randomization (MR)

To obtain valid instrumental variables (SNPs) for our analysis we assessed them against the three core assumptions for MR analysis: 1) That the SNPs were robustly associated with the exposure of interest. For that we obtained summary result statistics on genome-wide significant SNPs from our own GWAS. 2) That the SNPs were not associated with any known or unknown confounders. This is not an assumption that can be fully tested, however we used PhenoScanner ^72,73^ to assess whether any SNPs were associated with known confounders (described below). 3) That the SNPs were not associated with the outcomes through any other path than through the exposure. To test this assumption, we searched PhenoScanner ^72,73^ (detailed below) to see if our exposures of interest were associated with other potentially pleiotropic phenotypes.

### MR statistical analysis

We used the TwoSampleMR package ^74^ (https://github.com/MRCIEU/TwoSampleMR) in R version 4.2.2 (https://cran.r-project.org/). The outcome studies were obtained from https://www.magicinvestigators.org/ ^46^ and were external to the UK biobank. Specificially, we used the outcomes “fasting glucose adjusted for BMI” (mmol/L, n=200622), “2-hour post OGTT glucose adjusted for BMI” (mmol/L, n=63396), “fasting insulin adjusted for BMI” (pmol/L, n=151013) and “HbA1c” (%, n=146806) ^46^.

We performed a two-sample inverse variance weighted (IVW) analysis to assess the causal effect of CD300LG on metabolic factors (Supplementary Table 8 and 9). To explore potential violations of the core assumptions when using the full set of SNPs, we performed a heterogeneity test using Cochran’s Q, and a test for directional pleiotropy was conducted by assessing the degree to which the MR Egger intercept differed from zero ^75^. We also performed additional sensitivity analyses using MR Egger regression ^75^, weighted median ^76^, simple and weighted mode estimation methods ^77^. Effect estimates from the different sensitivity analysis were compared as a way of assessing the robustness of the results. To assess potential heterogeneity in the MR estimates we further performed MR-PRESSO ^46,78^ to detect (MR-PRESSO global test) and correct for horizontal pleiotropy via outlier removal (MR-PRESSO outlier test).

### Investigation of potentially pleiotropic SNPs

SNPs robustly associated with the exposure investigated in the MR analyses (serum CD300LG levels) were checked for other possible associations (PhenoScanner v2 ^72,73^, http://www.phenoscanner.medschl.cam.ac.uk/) which may contribute to a pleiotropic effect on the metabolic outcomes. Supplementary Table 10 lists the SNPs used in our analysis that many influence related phenotypes. Phenotypes from PhenoScanner were listed if they were associated with the SNPs or nearby variants in high LD (r^2^=0.8) at p-value level <1 x 10^-5^ and could have potential pleiotropic effects in the analysis.

### Data availability

mRNA sequencing data from MyoGlu can be found at https://exchmdpmg.medsch.ucla.edu/app/ as well as in GEO:GSE227419. Secretory proteins are available in the MetazSecKB data base at http://proteomics.ysu.edu/secretomes/animal/. The human serum proteomic NAFLD signature is available in the study of Govaere *et al* ^38^. Expression profiles in human liver cells are available in the Human Liver Cell Atlas ^39^. Data obtained from the UK biobank (Olink explore 1536 and measures of physical activity ^79^ can be found at https://biobank.ndph.ox.ac.uk/ukb/. Glucometabolic outcomes used in MR analyses are available at: https://www.magicinvestigators.org/ ^46^. Data from the GD-CAT database ^45^ is available from: https://pipeline.biochem.uci.edu/gtex/demo2/. Mice exercise data are available at https://motrpac-data.org/ and knock-out data at https://www.mousephenotype.org/. CD300LG expression values from a human tissue panel were obtained from Uhlén *et al.* ^42^. The single nuclei mRNA sequencing data from human adipose tissue was plotted in Seurat v. 4 by downloading processed data from the Single Cell Portal ^43^. The data can also be explored at: https://singlecell.broadinstitute.org/single_cell/study/SCP1376/a-single-cell-atlas-of-human-and-mouse-white-adipose-tissue). UK Biobank (https://www.ukbiobank.ac.uk/) data are available to researchers upon application to the individual cohorts via their websites. All other data used are publicly available and referenced according in the main text. For additional details and data inquiries, please contact Sindre Lee-Ødegård.

## Supporting information

SFigures

STables 1, 5-10

STables 2-4, 11

## Acknowledgements

South-Eastern Norway Regional Health Authority, Simon Fougners fund, Diabetesforbundet, Johan Selmer Kvanes’ legat til forskning og bekjempelse av sukkersyke. This research has been conducted using the UK Biobank resource (Reference 53641). DME is funded by an Australian National Health and Medical Research Council Investigator Grant (APP2017942). GHM is the recipient of an Australian Research Council Discovery Early Career Award (Project number: DE220101226) funded by the Australian Government and supported by the Research Council of Norway (Project grant: 325640 & Mobility grant: 287198). JKV is supported by The Medical Student Research Program at the University of Oslo. SL is supported by the Novo Nordisk Fonden Excellence Emerging Grant in Endocrinology and Metabolism 2023 (NNF23OC0082123).

## Author contributions

Conceptualization: SLØ, CAD, KIB. Methodology: SLØ, GHM, ED, DME. Data Collection: SLØ, FN. Data analysis: SLØ, TO, MH, GHM, ED. Visualization: SLØ, GHM and MH. Writing original draft: SLØ. Writing, review and editing: SLØ, MH, TO, GHM, ED, DME, JKV, HLG, FN, CAD, KIB. Supervision: CAD, KIB. Funding acquisition: SLØ, CAD, KIB. Project administration: KIB.

## References

1. Piercy, K.L., Troiano, R.P., Ballard, R.M., Carlson, S.A., Fulton, J.E., Galuska, D.A., George, S.M., and Olson, R.D. (2018). The Physical Activity Guidelines for Americans. Jama 320, 2020–2028. 10.1001/jama.2018.14854.

2. Hawley, J.A., and Lessard, S.J. (2008). Exercise training-induced improvements in insulin action. Acta Physiol (Oxf) 192, 127–135. 10.1111/j.1748-1716.2007.01783.x.

3. Bacchi, E., Negri, C., Zanolin, M.E., Milanese, C., Faccioli, N., Trombetta, M., Zoppini, G., Cevese, A., Bonadonna, R.C., Schena, F., et al. (2012). Metabolic effects of aerobic training and resistance training in type 2 diabetic subjects: a randomized controlled trial (the RAED2 study). Diabetes care 35, 676–682. 10.2337/dc11-1655.

4. Pedersen, B.K., Akerström, T.C., Nielsen, A.R., and Fischer, C.P. (2007). Role of myokines in exercise and metabolism. Journal of applied physiology (Bethesda, Md.: 1985) 103, 1093–1098. 10.1152/japplphysiol.00080.2007.

5. Chow, L.S., Gerszten, R.E., Taylor, J.M., Pedersen, B.K., van Praag, H., Trappe, S., Febbraio, M.A., Galis, Z.S., Gao, Y., Haus, J.M., et al. (2022). Exerkines in health, resilience and disease. Nat Rev Endocrinol 18, 273–289. 10.1038/s41574-022-00641-2.

6. Görgens, S.W., Eckardt, K., Jensen, J., Drevon, C.A., and Eckel, J. (2015). Exercise and Regulation of Adipokine and Myokine Production. Progress in molecular biology and translational science 135, 313–336. 10.1016/bs.pmbts.2015.07.002.

7. Lee-Ødegård, S., Olsen, T., Norheim, F., Drevon, C.A., and Birkeland, K.I. (2022). Potential Mechanisms for How Long-Term Physical Activity May Reduce Insulin Resistance. Metabolites 12. 10.3390/metabo12030208.

8. Pourteymour, S., Eckardt, K., Holen, T., Langleite, T., Lee, S., Jensen, J., Birkeland, K.I., Drevon, C.A., and Hjorth, M. (2017). Global mRNA sequencing of human skeletal muscle: Search for novel exercise-regulated myokines. Molecular metabolism 6, 352–365. 10.1016/j.molmet.2017.01.007.

9. Pedersen, B.K., and Febbraio, M.A. (2012). Muscles, exercise and obesity: skeletal muscle as a secretory organ. Nat Rev Endocrinol 8, 457–465. 10.1038/nrendo.2012.49.

10. Lee, S., Norheim, F., Langleite, T.M., Gulseth, H.L., Birkeland, K.I., and Drevon, C.A. (2019). Effects of long-term exercise on plasma adipokine levels and inflammation-related gene expression in subcutaneous adipose tissue in sedentary dysglycaemic, overweight men and sedentary normoglycaemic men of healthy weight. Diabetologia 62, 1048–1064. 10.1007/s00125-019-4866-5.

11. Bouassida, A., Chamari, K., Zaouali, M., Feki, Y., Zbidi, A., and Tabka, Z. (2010). Review on leptin and adiponectin responses and adaptations to acute and chronic exercise. Br J Sports Med 44, 620–630. 10.1136/bjsm.2008.046151.

12. Stanford, K.I., Lynes, M.D., Takahashi, H., Baer, L.A., Arts, P.J., May, F.J., Lehnig, A.C., Middelbeek, R.J.W., Richard, J.J., So, K., et al. (2018). 12,13-diHOME: An Exercise-Induced Lipokine that Increases Skeletal Muscle Fatty Acid Uptake. Cell metabolism 27, 1111–1120.e1113. 10.1016/j.cmet.2018.03.020.

13. Lee, S., Norheim, F., Gulseth, H.L., Langleite, T.M., Kolnes, K.J., Tangen, D.S., Stadheim, H.K., Gilfillan, G.D., Holen, T., Birkeland, K.I., et al. (2017). Interaction between plasma fetuin-A and free fatty acids predicts changes in insulin sensitivity in response to long-term exercise. Physiological reports 5. 10.14814/phy2.13183.

14. Kistner, T.M., Pedersen, B.K., and Lieberman, D.E. (2022). Interleukin 6 as an energy allocator in muscle tissue. Nature Metabolism 4, 170–179. 10.1038/s42255-022-00538-4.

15. Haugen, F., Norheim, F., Lian, H., Wensaas, A.J., Dueland, S., Berg, O., Funderud, A., Skålhegg, B.S., Raastad, T., and Drevon, C.A. (2010). IL-7 is expressed and secreted by human skeletal muscle cells. Am J Physiol Cell Physiol 298, C807–816. 10.1152/ajpcell.00094.2009.

16. Otaka, N., Shibata, R., Ohashi, K., Uemura, Y., Kambara, T., Enomoto, T., Ogawa, H., Ito, M., Kawanishi, H., Maruyama, S., et al. (2018). Myonectin Is an Exercise-Induced Myokine That Protects the Heart From Ischemia-Reperfusion Injury. Circ Res 123, 1326–1338. 10.1161/circresaha.118.313777.

17. Hjorth, M., Pourteymour, S., Görgens, S.W., Langleite, T.M., Lee, S., Holen, T., Gulseth, H.L., Birkeland, K.I., Jensen, J., Drevon, C.A., and Norheim, F. (2016). Myostatin in relation to physical activity and dysglycaemia and its effect on energy metabolism in human skeletal muscle cells. Acta Physiol (Oxf) 217, 45–60. 10.1111/apha.12631.

18. McPherron, A.C., Lawler, A.M., and Lee, S.J. (1997). Regulation of skeletal muscle mass in mice by a new TGF-beta superfamily member. Nature 387, 83–90. 10.1038/387083a0.

19. Rao, R.R., Long, J.Z., White, J.P., Svensson, K.J., Lou, J., Lokurkar, I., Jedrychowski, M.P., Ruas, J.L., Wrann, C.D., Lo, J.C., et al. (2014). Meteorin-like is a hormone that regulates immune-adipose interactions to increase beige fat thermogenesis. Cell 157, 1279–1291. 10.1016/j.cell.2014.03.065.

20. Kanzleiter, T., Rath, M., Gorgens, S.W., Jensen, J., Tangen, D.S., Kolnes, A.J., Kolnes, K.J., Lee, S., Eckel, J., Schurmann, A., and Eckardt, K. (2014). The myokine decorin is regulated by contraction and involved in muscle hypertrophy. Biochemical and biophysical research communications 450, 1089–1094. 10.1016/j.bbrc.2014.06.123.

21. Malin, S.K., del Rincon, J.P., Huang, H., and Kirwan, J.P. (2014). Exercise-induced lowering of fetuin-A may increase hepatic insulin sensitivity. Medicine and science in sports and exercise 46, 2085–2090. 10.1249/mss.0000000000000338.

22. Catoire, M., Alex, S., Paraskevopulos, N., Mattijssen, F., Evers-van Gogh, I., Schaart, G., Jeppesen, J., Kneppers, A., Mensink, M., Voshol, P.J., et al. (2014). Fatty acid-inducible ANGPTL4 governs lipid metabolic response to exercise. Proceedings of the National Academy of Sciences of the United States of America 111, E1043–1052. 10.1073/pnas.1400889111.

23. Norheim, F., Hjorth, M., Langleite, T.M., Lee, S., Holen, T., Bindesboll, C., Stadheim, H.K., Gulseth, H.L., Birkeland, K.I., Kielland, A., et al. (2014). Regulation of angiopoietin-like protein 4 production during and after exercise. Physiological reports 2. 10.14814/phy2.12109.

24. Contrepois, K., Wu, S., Moneghetti, K.J., Hornburg, D., Ahadi, S., Tsai, M.S., Metwally, A.A., Wei, E., Lee-McMullen, B., Quijada, J.V., et al. (2020). Molecular Choreography of Acute Exercise. Cell 181, 1112–1130.e1116. 10.1016/j.cell.2020.04.043.

25. Diaz-Canestro, C., Chen, J., Liu, Y., Han, H., Wang, Y., Honoré, E., Lee, C.H., Lam, K.S.L., Tse, M.A., and Xu, A. (2023). A machine-learning algorithm integrating baseline serum proteomic signatures predicts exercise responsiveness in overweight males with prediabetes. Cell Rep Med 4, 100944. 10.1016/j.xcrm.2023.100944.

26. Robbins, J.M., Peterson, B., Schranner, D., Tahir, U.A., Rienmüller, T., Deng, S., Keyes, M.J., Katz, D.H., Beltran, P.M.J., Barber, J.L., et al. (2021). Human plasma proteomic profiles indicative of cardiorespiratory fitness. Nat Metab 3, 786–797. 10.1038/s42255-021-00400-z.

27. Robbins, J.M., Rao, P., Deng, S., Keyes, M.J., Tahir, U.A., Katz, D.H., Beltran, P.M.J., Marchildon, F., Barber, J.L., Peterson, B., et al. (2023). Plasma proteomic changes in response to exercise training are associated with cardiorespiratory fitness adaptations. JCI Insight 8. 10.1172/jci.insight.165867.

28. Mi, M.Y., Barber, J.L., Rao, P., Farrell, L.A., Sarzynski, M.A., Bouchard, C., Robbins, J.M., and Gerszten, R.E. (2023). Plasma Proteomic Kinetics in Response to Acute Exercise. Mol Cell Proteomics 22, 100601. 10.1016/j.mcpro.2023.100601.

29. Langleite, T.M., Jensen, J., Norheim, F., Gulseth, H.L., Tangen, D.S., Kolnes, K.J., Heck, A., Storas, T., Grothe, G., Dahl, M.A., et al. (2016). Insulin sensitivity, body composition and adipose depots following 12 w combined endurance and strength training in dysglycemic and normoglycemic sedentary men. Archives of physiology and biochemistry 122, 167–179. 10.1080/13813455.2016.1202985.

30. Sudlow, C., Gallacher, J., Allen, N., Beral, V., Burton, P., Danesh, J., Downey, P., Elliott, P., Green, J., Landray, M., et al. (2015). UK Biobank: An Open Access Resource for Identifying the Causes of a Wide Range of Complex Diseases of Middle and Old Age. PLOS Medicine 12, e1001779. 10.1371/journal.pmed.1001779.

31. Hamaguchi, H., Dohi, K., Sakai, T., Taoka, M., Isobe, T., Matsui, T.S., Deguchi, S., Furuichi, Y., Fujii, N.L., and Manabe, Y. (2023). PDGF-B secreted from skeletal muscle enhances myoblast proliferation and myotube maturation via activation of the PDGFR signaling cascade. Biochemical and biophysical research communications 639, 169–175. 10.1016/j.bbrc.2022.11.085.

32. Lories, R.J., Peeters, J., Szlufcik, K., Hespel, P., and Luyten, F.P. (2009). Deletion of frizzled-related protein reduces voluntary running exercise performance in mice. Osteoarthritis Cartilage 17, 390–396. 10.1016/j.joca.2008.07.018.

33. Casas-Fraile, L., Cornelis, F.M., Costamagna, D., Rico, A., Duelen, R., Sampaolesi, M.M., López de Munain, A., Lories, R.J., and Sáenz, A. (2020). Frizzled related protein deficiency impairs muscle strength, gait and calpain 3 levels. Orphanet Journal of Rare Diseases 15, 119. 10.1186/s13023-020-01372-1.

34. López-Bellón, S., Rodríguez-López, S., González-Reyes, J.A., Burón, M.I., de Cabo, R., and Villalba, J.M. (2022). CYB5R3 overexpression preserves skeletal muscle mitochondria and autophagic signaling in aged transgenic mice. Geroscience 44, 2223–2241. 10.1007/s11357-022-00574-8.

35. Tassi, E., Garman, K.A., Schmidt, M.O., Ma, X., Kabbara, K.W., Uren, A., Tomita, Y., Goetz, R., Mohammadi, M., Wilcox, C.S., et al. (2018). Fibroblast Growth Factor Binding Protein 3 (FGFBP3) impacts carbohydrate and lipid metabolism. Sci Rep 8, 15973. 10.1038/s41598-018-34238-5.

36. Qian, Q., Hu, F., Yu, W., Leng, D., Li, Y., Shi, H., Deng, D., Ding, K., Liang, C., and Liu, J. (2023). SWAP70 Overexpression Protects Against Pathological Cardiac Hypertrophy in a TAK1-Dependent Manner. J Am Heart Assoc 12, e028628. 10.1161/jaha.122.028628.

37. Chen, H.H., Luche, R., Wei, B., and Tonks, N.K. (2004). Characterization of two distinct dual specificity phosphatases encoded in alternative open reading frames of a single gene located on human chromosome 10q22.2. The Journal of biological chemistry 279, 41404–41413. 10.1074/jbc.M405286200.

38. Govaere, O., Hasoon, M., Alexander, L., Cockell, S., Tiniakos, D., Ekstedt, M., Schattenberg, J.M., Boursier, J., Bugianesi, E., Ratziu, V., et al. (2023). A proteo-transcriptomic map of non-alcoholic fatty liver disease signatures. Nature Metabolism 5, 572–578. 10.1038/s42255-023-00775-1.

39. Aizarani, N., Saviano, A., Sagar, Mailly, L., Durand, S., Herman, J.S., Pessaux, P., Baumert, T.F., and Grün, D. (2019). A human liver cell atlas reveals heterogeneity and epithelial progenitors. Nature 572, 199–204. 10.1038/s41586-019-1373-2.

40. Montgomery, M.K., Bayliss, J., Devereux, C., Bezawork-Geleta, A., Roberts, D., Huang, C., Schittenhelm, R.B., Ryan, A., Townley, S.L., Selth, L.A., et al. (2020). SMOC1 is a glucose-responsive hepatokine and therapeutic target for glycemic control. Sci Transl Med 12. 10.1126/scitranslmed.aaz8048.

41. Ghodsian, N., Gagnon, E., Bourgault, J., Gobeil, É., Manikpurage, H.D., Perrot, N., Girard, A., Mitchell, P.L., and Arsenault, B.J. (2021). Blood Levels of the SMOC1 Hepatokine Are Not Causally Linked with Type 2 Diabetes: A Bidirectional Mendelian Randomization Study. Nutrients 13. 10.3390/nu13124208.

42. Uhlén, M., Fagerberg, L., Hallström, B.M., Lindskog, C., Oksvold, P., Mardinoglu, A., Sivertsson, Å., Kampf, C., Sjöstedt, E., Asplund, A., et al. (2015). Proteomics. Tissue-based map of the human proteome. Science (New York, N.Y.) 347, 1260419. 10.1126/science.1260419.

43. Emont, M.P., Jacobs, C., Essene, A.L., Pant, D., Tenen, D., Colleluori, G., Di Vincenzo, A., Jørgensen, A.M., Dashti, H., Stefek, A., et al. (2022). A single-cell atlas of human and mouse white adipose tissue. Nature 603, 926–933. 10.1038/s41586-022-04518-2.

44. Battle, A., Brown, C.D., Engelhardt, B.E., and Montgomery, S.B. (2017). Genetic effects on gene expression across human tissues. Nature 550, 204–213. 10.1038/nature24277.

45. Zhou, M., Tamburini, I., Van, C., Molendijk, J., Nguyen, C.M., Chang, I.Y.-Y., Johnson, C., Velez, L.M., Cheon, Y., Yeo, R., et al. (2024). Leveraging inter-individual transcriptional correlation structure to infer discrete signaling mechanisms across metabolic tissues. eLife 12, RP88863. 10.7554/eLife.88863.

46. Chen, J., Spracklen, C.N., Marenne, G., Varshney, A., Corbin, L.J., Luan, J., Willems, S.M., Wu, Y., Zhang, X., Horikoshi, M., et al. (2021). The trans-ancestral genomic architecture of glycemic traits. Nat Genet 53, 840–860. 10.1038/s41588-021-00852-9.

47. Sanford, J.A., Nogiec, C.D., Lindholm, M.E., Adkins, J.N., Amar, D., Dasari, S., Drugan, J.K., Fernández, F.M., Radom-Aizik, S., Schenk, S., et al. (2020). Molecular Transducers of Physical Activity Consortium (MoTrPAC): Mapping the Dynamic Responses to Exercise. Cell 181, 1464–1474. 10.1016/j.cell.2020.06.004.

48. Dickinson, M.E., Flenniken, A.M., Ji, X., Teboul, L., Wong, M.D., White, J.K., Meehan, T.F., Weninger, W.J., Westerberg, H., Adissu, H., et al. (2016). High-throughput discovery of novel developmental phenotypes. Nature 537, 508–514. 10.1038/nature19356.

49. Takatsu, H., Hase, K., Ohmae, M., Ohshima, S., Hashimoto, K., Taniura, N., Yamamoto, A., and Ohno, H. (2006). CD300 antigen like family member G: A novel Ig receptor like protein exclusively expressed on capillary endothelium. Biochemical and biophysical research communications 348, 183–191. 10.1016/j.bbrc.2006.07.047.

50. Liang, X., Yee, S.W., Chien, H.C., Chen, E.C., Luo, Q., Zou, L., Piao, M., Mifune, A., Chen, L., Calvert, M.E., et al. (2018). Organic cation transporter 1 (OCT1) modulates multiple cardiometabolic traits through effects on hepatic thiamine content. PLoS Biol 16, e2002907. 10.1371/journal.pbio.2002907.

51. Chen, L., Shu, Y., Liang, X., Chen, E.C., Yee, S.W., Zur, A.A., Li, S., Xu, L., Keshari, K.R., Lin, M.J., et al. (2014). OCT1 is a high-capacity thiamine transporter that regulates hepatic steatosis and is a target of metformin. Proceedings of the National Academy of Sciences of the United States of America 111, 9983–9988. 10.1073/pnas.1314939111.

52. Jacob, Y., Anderton, R.S., Cochrane Wilkie, J.L., Rogalski, B., Laws, S.M., Jones, A., Spiteri, T., Hince, D., and Hart, N.H. (2022). Genetic Variants within NOGGIN, COL1A1, COL5A1, and IGF2 are Associated with Musculoskeletal Injuries in Elite Male Australian Football League Players: A Preliminary Study. Sports Medicine – Open 8, 126. 10.1186/s40798-022-00522-y.

53. Barry, J.C., Simtchouk, S., Durrer, C., Jung, M.E., and Little, J.P. (2017). Short-Term Exercise Training Alters Leukocyte Chemokine Receptors in Obese Adults. Medicine and science in sports and exercise 49, 1631–1640. 10.1249/mss.0000000000001261.

54. Meinken, J., Walker, G., Cooper, C.R., and Min, X.J. (2015). MetazSecKB: the human and animal secretome and subcellular proteome knowledgebase. Database (Oxford) 2015. 10.1093/database/bav077.

55. Umemoto, E., Takeda, A., Jin, S., Luo, Z., Nakahogi, N., Hayasaka, H., Lee, C.M., Tanaka, T., and Miyasaka, M. (2013). Dynamic changes in endothelial cell adhesion molecule nepmucin/CD300LG expression under physiological and pathological conditions. PloS one 8, e83681. 10.1371/journal.pone.0083681.

56. Støy, J., Grarup, N., Hørlyck, A., Ibsen, L., Rungby, J., Poulsen, P.L., Brandslund, I., Christensen, C., Hansen, T., Pedersen, O., et al. (2014). Blood pressure levels in male carriers of Arg82Cys in CD300LG. PloS one 9, e109646. 10.1371/journal.pone.0109646.

57. Umemoto, E., Tanaka, T., Kanda, H., Jin, S., Tohya, K., Otani, K., Matsutani, T., Matsumoto, M., Ebisuno, Y., Jang, M.H., et al. (2006). Nepmucin, a novel HEV sialomucin, mediates L-selectin-dependent lymphocyte rolling and promotes lymphocyte adhesion under flow. J Exp Med 203, 1603–1614. 10.1084/jem.20052543.

58. Surakka, I., Horikoshi, M., Mägi, R., Sarin, A.-P., Mahajan, A., Lagou, V., Marullo, L., Ferreira, T., Miraglio, B., Timonen, S., et al. (2015). The impact of low-frequency and rare variants on lipid levels. Nature Genetics 47, 589–597. 10.1038/ng.3300.

59. Støy, J., Kampmann, U., Mengel, A., Magnusson, N.E., Jessen, N., Grarup, N., Rungby, J., Stødkilde-Jørgensen, H., Brandslund, I., Christensen, C., et al. (2015). Reduced CD300LG mRNA tissue expression, increased intramyocellular lipid content and impaired glucose metabolism in healthy male carriers of Arg82Cys in CD300LG: a novel genometabolic cross-link between CD300LG and common metabolic phenotypes. BMJ Open Diabetes Res Care 3, e000095. 10.1136/bmjdrc-2015-000095.

60. Takatsu, H., Hase, K., Ohmae, M., Ohshima, S., Hashimoto, K., Taniura, N., Yamamoto, A., and Ohno, H. (2006). CD300 antigen like family member G: A novel Ig receptor like protein exclusively expressed on capillary endothelium. Biochemical and biophysical research communications 348, 183–191. 10.1016/j.bbrc.2006.07.047.

61. Van Pelt, D.W., Guth, L.M., and Horowitz, J.F. (2017). Aerobic exercise elevates markers of angiogenesis and macrophage IL-6 gene expression in the subcutaneous adipose tissue of overweight-to-obese adults. Journal of applied physiology 123, 1150–1159. 10.1152/japplphysiol.00614.2017.

62. Ross, M., Kargl, C.K., Ferguson, R., Gavin, T.P., and Hellsten, Y. (2023). Exercise-induced skeletal muscle angiogenesis: impact of age, sex, angiocrines and cellular mediators. European Journal of Applied Physiology 123, 1415–1432. 10.1007/s00421-022-05128-6.

63. Lee, S., Gulseth, H.L., Langleite, T.M., Norheim, F., Olsen, T., Refsum, H., Jensen, J., Birkeland, K.I., and Drevon, C.A. (2021). Branched-chain amino acid metabolism, insulin sensitivity and liver fat response to exercise training in sedentary dysglycaemic and normoglycaemic men. Diabetologia 64, 410–423. 10.1007/s00125-020-05296-0.

64. Sudlow, C., Gallacher, J., Allen, N., Beral, V., Burton, P., Danesh, J., Downey, P., Elliott, P., Green, J., Landray, M., et al. (2015). UK biobank: an open access resource for identifying the causes of a wide range of complex diseases of middle and old age. PLoS Med 12, e1001779. 10.1371/journal.pmed.1001779.

65. Bycroft, C., Freeman, C., Petkova, D., Band, G., Elliott, L.T., Sharp, K., Motyer, A., Vukcevic, D., Delaneau, O., O’Connell, J., et al. (2018). The UK Biobank resource with deep phenotyping and genomic data. Nature 562, 203–209. 10.1038/s41586-018-0579-z.

66. Auton, A., Brooks, L.D., Durbin, R.M., Garrison, E.P., Kang, H.M., Korbel, J.O., Marchini, J.L., McCarthy, S., McVean, G.A., and Abecasis, G.R. (2015). A global reference for human genetic variation. Nature 526, 68–74. 10.1038/nature15393.

67. Loh, P.-R., Tucker, G., Bulik-Sullivan, B.K., Vilhjálmsson, B.J., Finucane, H.K., Salem, R.M., Chasman, D.I., Ridker, P.M., Neale, B.M., Berger, B., et al. (2015). Efficient Bayesian mixed-model analysis increases association power in large cohorts. Nature Genetics 47, 284–290. 10.1038/ng.3190.

68. Wu, Y., Burch, K.S., Ganna, A., Pajukanta, P., Pasaniuc, B., and Sankararaman, S. (2022). Fast estimation of genetic correlation for biobank-scale data. The American Journal of Human Genetics 109, 24–32. 10.1016/j.ajhg.2021.11.015.

69. Lee, J.J., McGue, M., Iacono, W.G., and Chow, C.C. (2018). The accuracy of LD Score regression as an estimator of confounding and genetic correlations in genome-wide association studies. Genet Epidemiol 42, 783–795. 10.1002/gepi.22161.

70. Wang, K., Li, M., and Hakonarson, H. (2010). ANNOVAR: functional annotation of genetic variants from high-throughput sequencing data. Nucleic Acids Res 38, e164. 10.1093/nar/gkq603.

71. Purcell, S., Neale, B., Todd-Brown, K., Thomas, L., Ferreira, M.A., Bender, D., Maller, J., Sklar, P., de Bakker, P.I., Daly, M.J., and Sham, P.C. (2007). PLINK: a tool set for whole-genome association and population-based linkage analyses. American journal of human genetics 81, 559–575. 10.1086/519795.

72. Staley, J.R., Blackshaw, J., Kamat, M.A., Ellis, S., Surendran, P., Sun, B.B., Paul, D.S., Freitag, D., Burgess, S., Danesh, J., et al. (2016). PhenoScanner: a database of human genotype-phenotype associations. Bioinformatics (Oxford, England) 32, 3207–3209. 10.1093/bioinformatics/btw373.

73. Kamat, M.A., Blackshaw, J.A., Young, R., Surendran, P., Burgess, S., Danesh, J., Butterworth, A.S., and Staley, J.R. (2019). PhenoScanner V2: an expanded tool for searching human genotype-phenotype associations. Bioinformatics (Oxford, England) 35, 4851–4853. 10.1093/bioinformatics/btz469.

74. Hemani, G., Zheng, J., Elsworth, B., Wade, K.H., Haberland, V., Baird, D., Laurin, C., Burgess, S., Bowden, J., Langdon, R., et al. (2018). The MR-Base platform supports systematic causal inference across the human phenome. eLife 7, e34408. 10.7554/eLife.34408.

75. Bowden, J., Davey Smith, G., and Burgess, S. (2015). Mendelian randomization with invalid instruments: effect estimation and bias detection through Egger regression. Int J Epidemiol 44, 512–525. 10.1093/ije/dyv080.

76. Bowden, J., Del Greco, M.F., Minelli, C., Davey Smith, G., Sheehan, N.A., and Thompson, J.R. (2016). Assessing the suitability of summary data for two-sample Mendelian randomization analyses using MR-Egger regression: the role of the I2 statistic. Int J Epidemiol 45, 1961–1974. 10.1093/ije/dyw220.

77. Hartwig, F.P., Davey Smith, G., and Bowden, J. (2017). Robust inference in summary data Mendelian randomization via the zero modal pleiotropy assumption. Int J Epidemiol 46, 1985–1998. 10.1093/ije/dyx102.

78. Verbanck, M., Chen, C.-Y., Neale, B., and Do, R. (2018). Detection of widespread horizontal pleiotropy in causal relationships inferred from Mendelian randomization between complex traits and diseases. Nature Genetics 50, 693–698. 10.1038/s41588-018-0099-7.

79. Sophie, C., Josephine, Y.C., Michael, C., Adrian, B., and Michael, I.T. (2016). Cross-sectional study of diet, physical activity, television viewing and sleep duration in 233 110 adults from the UK Biobank; the behavioural phenotype of cardiovascular disease and type 2 diabetes. BMJ Open 6, e010038. 10.1136/bmjopen-2015-010038.

