## Supplementary material for "Serum proteomic profiling of physical activity reveals CD300LG as a novel exerkine with a potential causal link to glucose homeostasis": SFigures

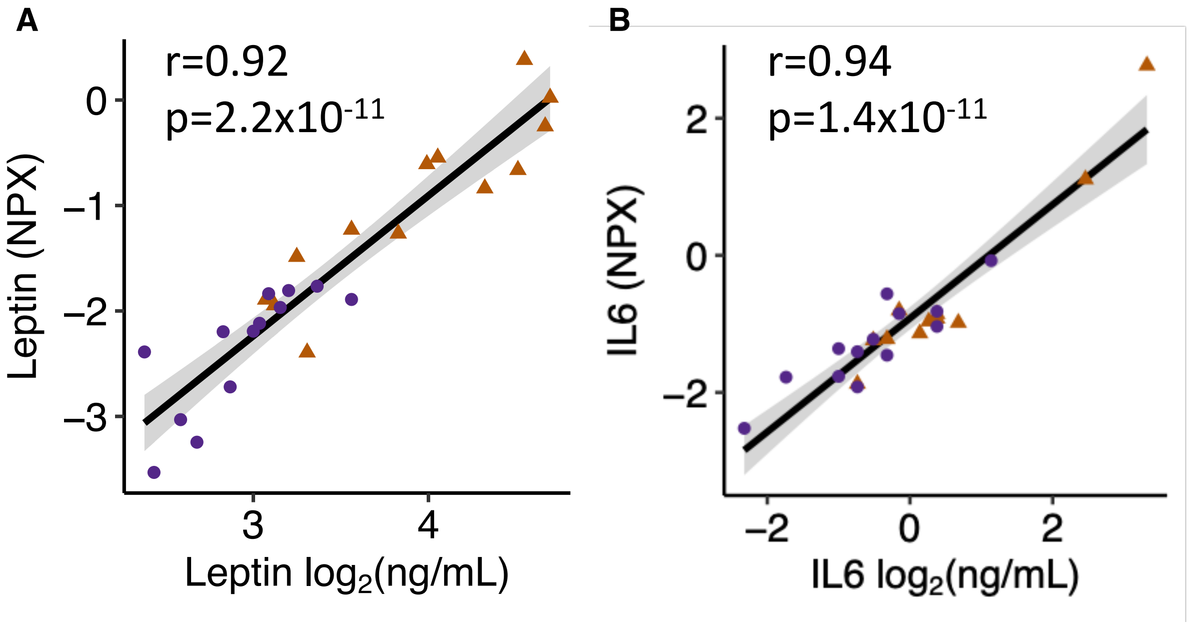


**Supplementary Figure 1.** Olink vs. ELISA for (A) serum leptin and (B) IL6 protein levels.

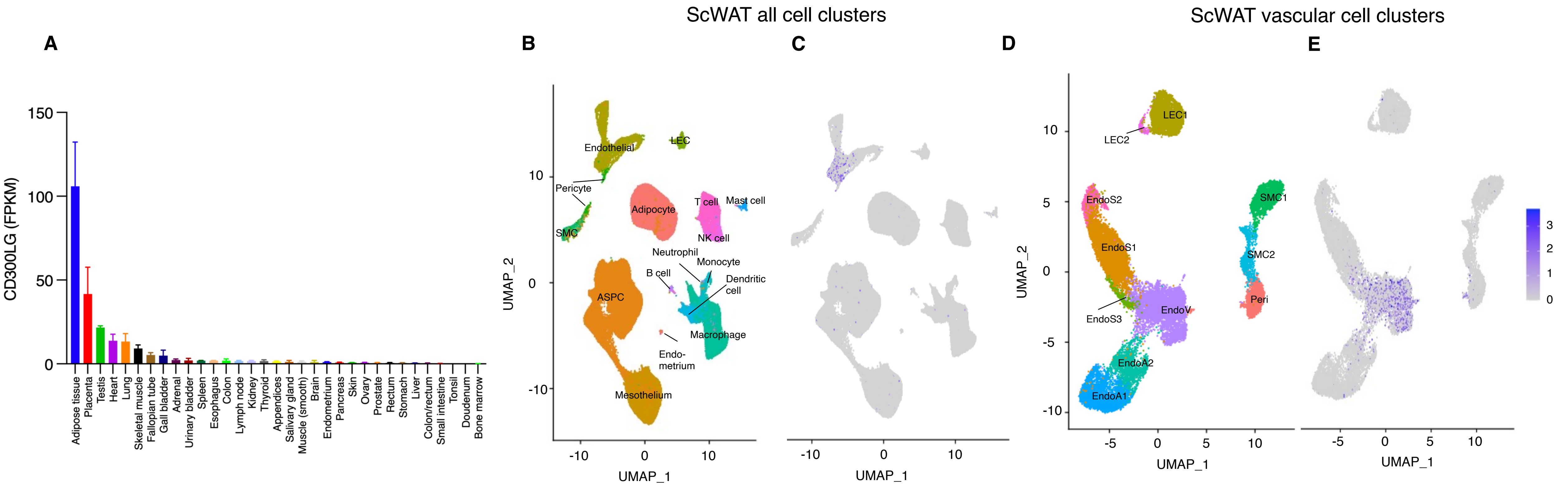


**Supplementary Figure 2. Tissue and cell specific expression of CD300LG.** (A) mRNA levels of CD300LG in a human tissue panel (see Methods). (B-C) snRNAseq of human adipose tissue, displaying (B) all detected cell clusters and (C) CD300LG related to the clusters (purple color). (D-E) Similar to B-C, but showing the (D) vascular cell clusters and (E) the corresponding expression of CD300LG (purple color). FPKM = Fragments per kilobase of transcript per million mapped reads. Data were obtained from Uhlen et al. [42] and from Emont et al. [43] and can be explored at https://singlecell.broadinstitute.org/single_cell

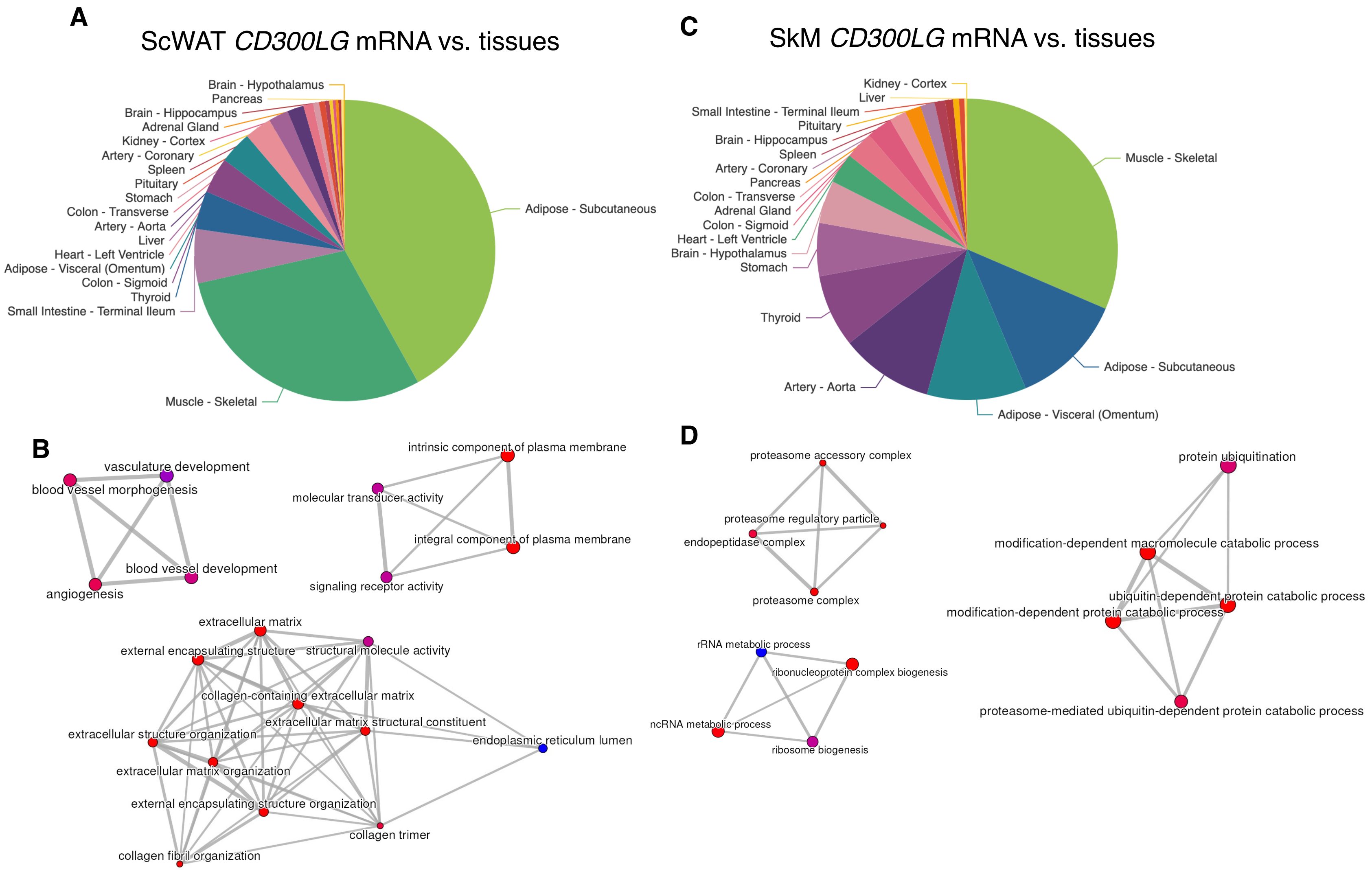


**Supplementary Figure 3. *CD300LG* mRNA correlations in men.** (A) Subcutanous white adipose tissue (ScWAT) *CD300LG* mRNA and correlations with tissue gene expression. (B) The top 3 networks of *CD300LG*-related biological processes in adipose tissue. (C) Skeletal muscle (SkM) *CD300LG* mRNA and correlations with tissue gene expression. (B) The top 3 networks of *CD300LG*-related biological processes in skeletal muscle. Data were obtained from the GD-CAT (Genetically-Derived Correlations Across Tissues) database described in Battle et *a. l*[44] and Zhou et *al.* [45].

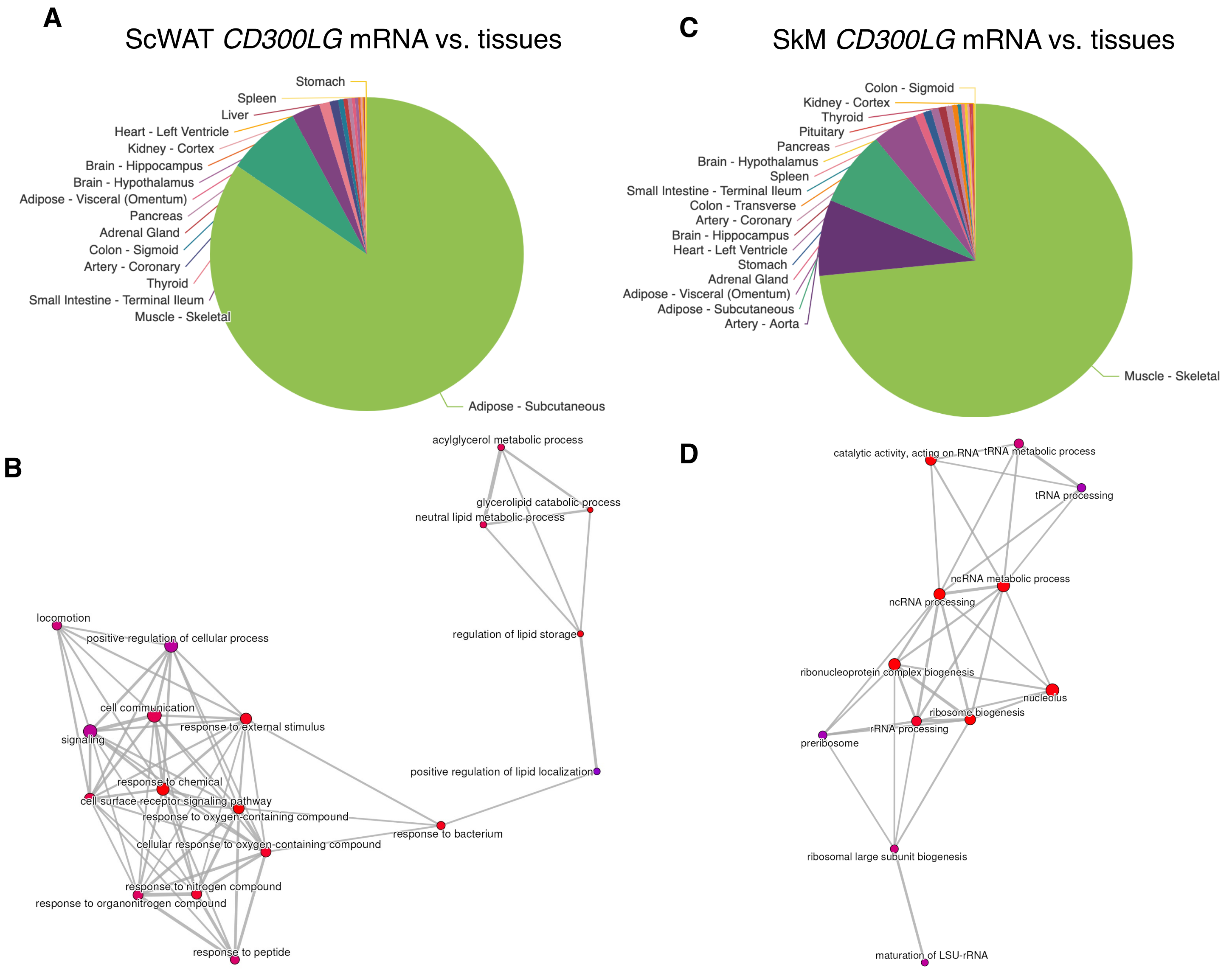


**Supplementary Figure 4. *CD300LG* mRNA correlations in women.** (A) Subcutaneous white adipose tissue (ScWAT) *CD300LG* mRNA and correlations with tissue gene expression. (B) The top network of *CD300LG*-related biological processes in adipose tissue. (C) Skeletal muscle (SkM) *CD300LG* mRNA and correlations with tissue gene expression. (B) The top network of *CD300LG*-related biological processes in skeletal muscle. Data were obtained from the GD-CAT (Genetically-Derived Correlations Across Tissues) database described in Battle et *al.* [44] and Zhou et *al.* [45].

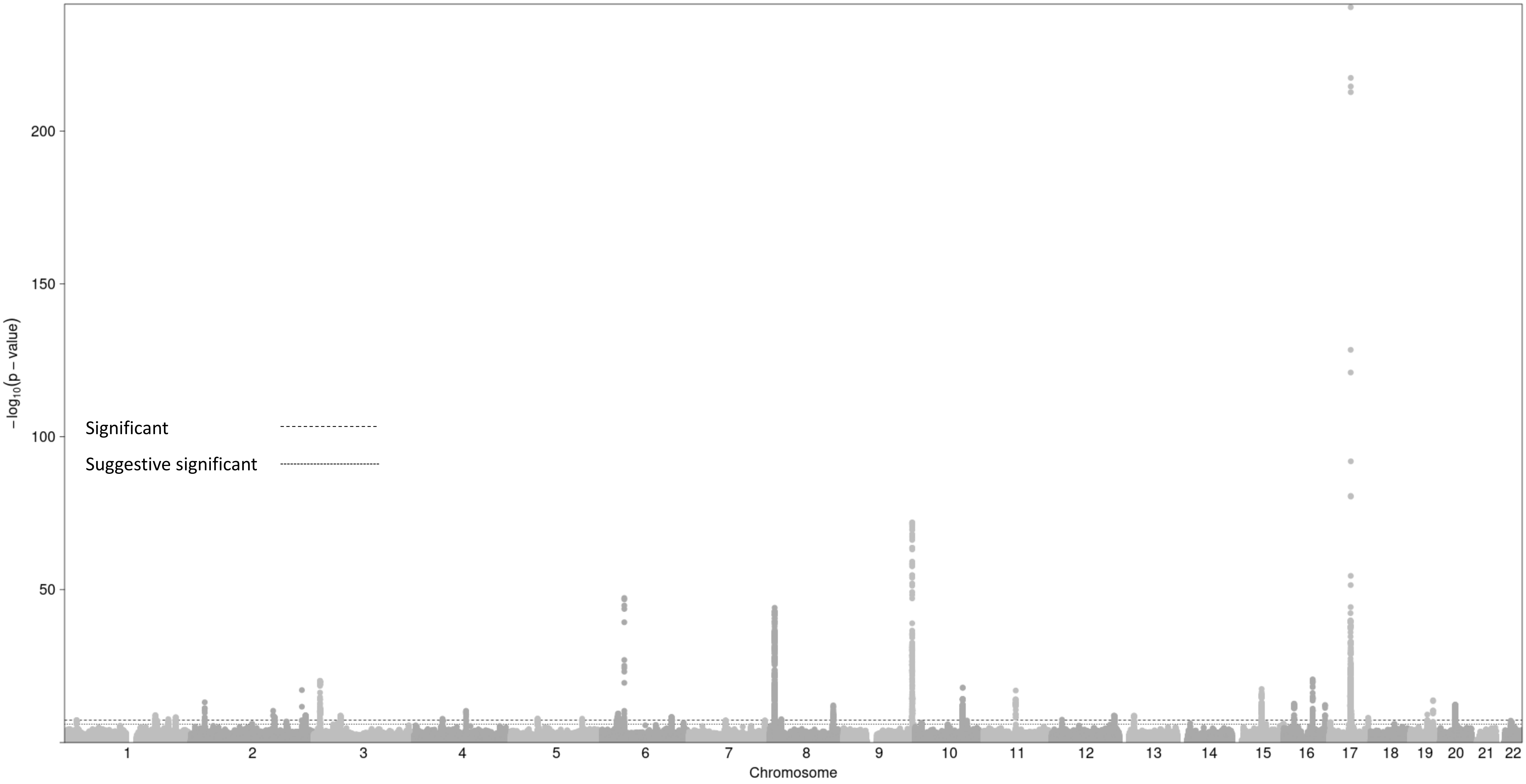


**Supplementary Figure 5.** Manhattan plot for serum CD300LG protein levels GWAS. The dashed line denotes ‘genome-wide significance’ threshold of $p<5\times10^{-8}$ whilst the dotted line denotes ‘suggestive significance’ threshold of $p<1\times10^{-6}$.

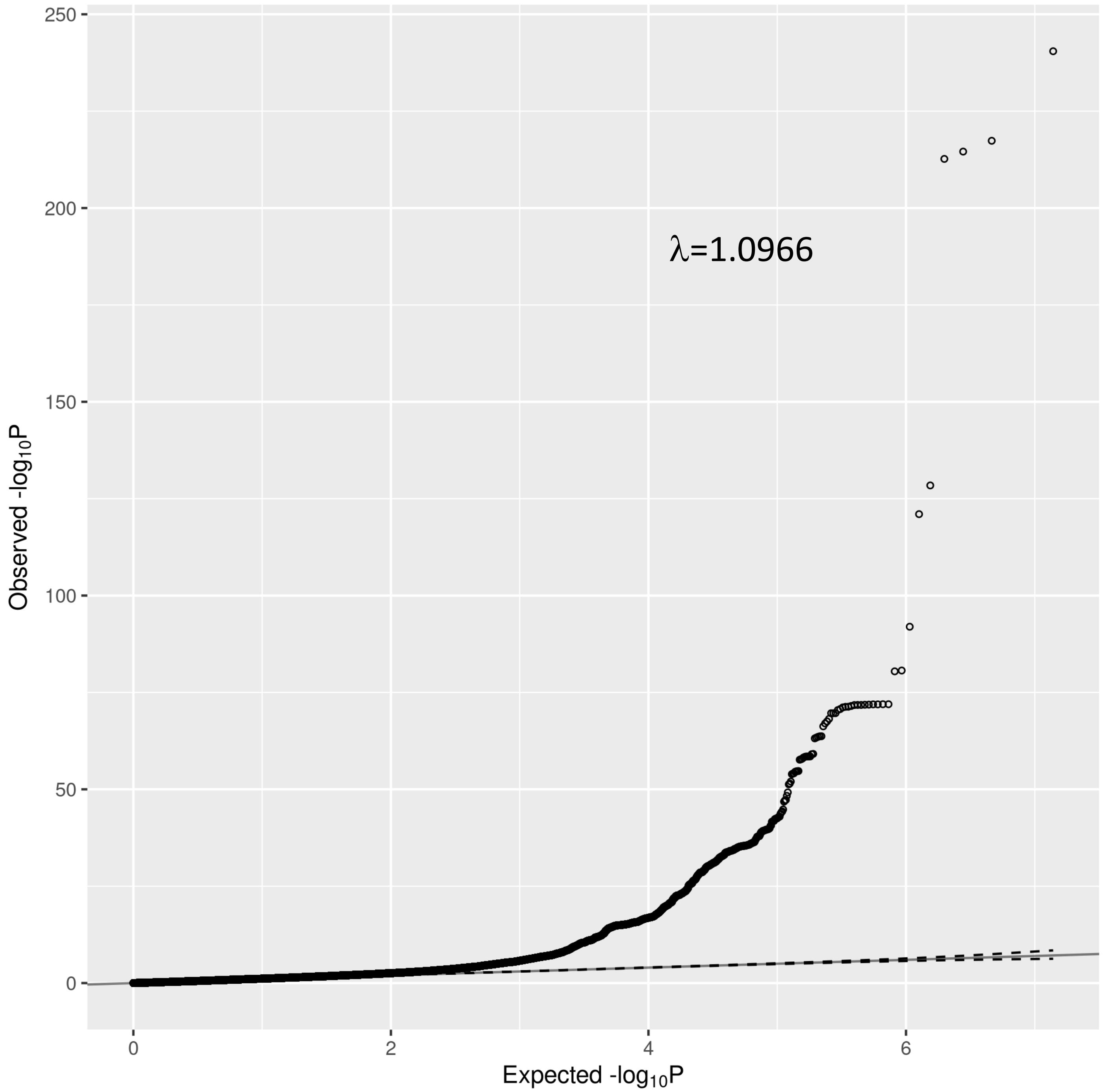


**Supplementary Figure 6.** Quantile-Quantile plot for CD300LG protein GWAS. Comparison of observed and expected -$\log_{10}$ *p*-values expected under the null distribution of no genetic association across the genome. The dotted line represent the pointwise 95% confidence interval expected under the null hypothesis of no association. The genomic inflation factor (lambda=1.0966) and LD score intercept (1.039) were consistent with our GWAS being well controlled for population stratification and other possible biases

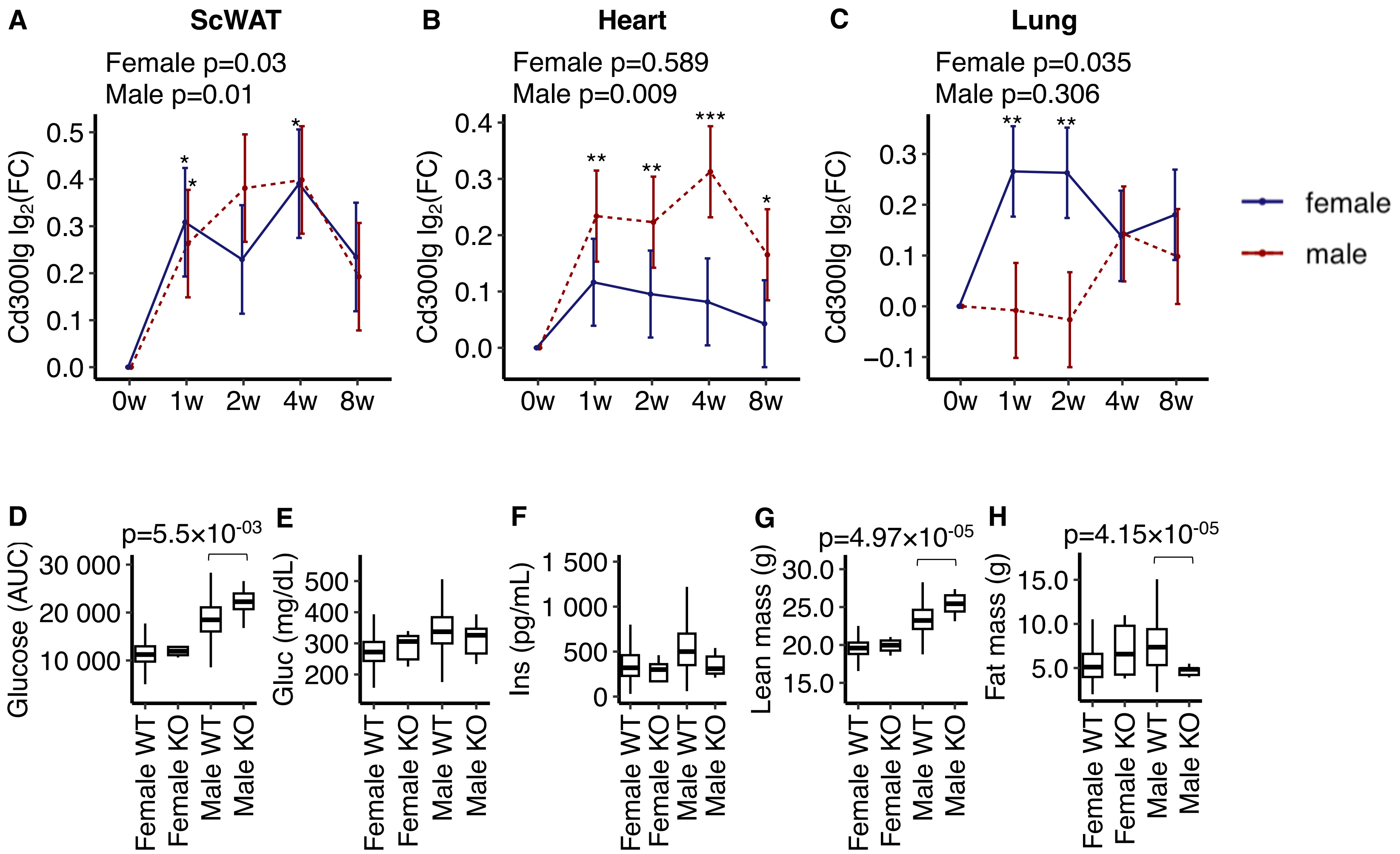


**Supplementary Figure 7.** MoTrPAC mice data (n=12-15 female and n=12-15 male mice) from Sanford et *al.* [47] showing Cd300lg protein levels in (A) subcutaneous white adipose tissue (scWAT), (B) heart muscle and (C) lung tissue in response to endurance exercise for 8 weeks. International Mouse Phenotyping Consortium data obtained from Dickinson et *al.*[48] showing (D) effects of *Cd300lg* on glucose tolerance, (E) fasting glucose concentration, (F) fasting insulin concentration, (G) lean body mass, and (H) body fat mass. Mice included wild type (n=1516 females and n=1520 males), and n=7 females and n=7 males knock out mice. Data were analyzed using mixed linear regression. *p<0.05, **p<0.01 and ***p<0.001. AUC = areal under the curve. w = week. WT = wild type. KO = knock out.
