## Supplementary material for "Serum proteomic profiling of physical activity reveals CD300LG as a novel exerkine with a potential causal link to glucose homeostasis": STables 1, 5-10

*Supplementary Table 1. Subject characteristics at baseline and changes observed after 12 weeks of exercise.*

| | **Baseline** | | **%-change** | |
|---|---|---|---|---|
| | NW (*n* = 13) | OW (*n* = 13) | NW (*n* = 13) | OW (*n* = 13) |
| Age (years) | 50 (7) | 53 (6) | | |
| **Body composition** | | | | |
| Weight (kg) | 78.5 (8.2) | 95.4 (10.2) * | −0.3 (2.1) | −1.7 (2.4) † |
| BMI (kg/m$^2$) | 23.5 (2.0) | 29.0 (2.4) * | 0.0 (2.0) | −1.2 (4.5) |
| FFM (kg) [a] | 34.9 (3.5) | 37.7 (5.0) | 6.4 (3.8) † | 5.3 (2.7) † |
| ScWAT (kg) [a] | 10.3 (2.7) | 18.0 (4.2) * | −6.6 (9.2) † | −7.3 (6.0) † |
| IAAT (kg) [a] | 4.0 (2.0) | 8.8 (2.6) * | −16.9 (15.1) † | −19.4 (10.8) † |
| Hepatic fat (AU) [b] | 2.8 (2.2) | 9.1 (5.9) * | −23.3 (50.7) † | −27.4 (15.8) † |
| Thigh muscle area (AU) [a] | 20.3 (2.9) | 24.4 (3.1) * | 9.7 (4.7) † | 7.1 (6.7) † |
| **Physical fitness** | | | | |
| $VO_2$max (ml/kg/min) | 44.1 (4.4) | 37.1 (4.9) * | 13.2 (9.7) † | 13.3 (7.7) † |
| Chest press (kg) | 65.6 (16.8) | 68.7 (13.7) | 18.4 (8.7) † | 13.6 (8.4) † |
| Pull down (kg) | 68.8 (9.3) | 75.6 (15.1) | 18.3 (10.1) † | 13.7 (7.3) † |
| Leg press (kg) | 199.6 (36.9) | 248.7 (30.3) * | 9.8 (7.6) † | 12.5 (8.4) † |
| **Glucose metabolism** | | | | |
| HbA1c (mmol/mol) | 33 (4) | 37 (4) * | N.A. | N.A. |
| HbA1c (%) | 5.2 (0.2) | 5.5 (0.4) * | N.A. | N.A. |
| Glucose (mmol/l) | 5.4 (0.5) | 5.9 (0.3) * | 3.1 (4.5) † | 1.8 (6.8) |
| C-Peptide (pmol/l) | 588.0 (117.8) | 932.8 (248.9) * | 7.3 (23.8) | 12.3 (45.3) |
| Insulin (pmol/l) | 38.5 (18.6) | 65.3 (27.1) * | 15.1 (49.2) | 27.6 (66.2) |
| GIR (mg/kg/min) | 7.6 (1.6) | 4.2 (1.8) * | 37.8 (30.1) † | 44.4 (58.8) † |

**Notes:** [a] *n* = 12 control, [b] *n* = 10 control, and *n* = 9 OW. * $p < 0.05$ between groups (OW vs. NW), and † $p < 0.05$ baseline vs. 12 w within group. Between groups comparison were performed using unpaired *t*-tests, and within group comparisons were performed using paired *t*-tests. Logarithmic transformation was performed to approximate normal distribution, if necessary, and back transformed for presentation. Data represent means (SD).

**Abbreviations:** NW = normal weight. OW = overweight. N.A. = not available, AU = arbitrary units, BMI = body mass index, FFM = fat free mass, AT = adipose tissue, Sc = subcutaneous, IA = intra-abdominal, GIR = glucose infusion rate.

*Supplementary Table 5: Descriptive statistics UK biobank*

| | **All individuals (n=52,981)** | | **Women (n=28,579)** | | **Men (n=24,402)** | | |
|---|---|---|---|---|---|---|---|
| | Mean | SD | Mean | SD | Mean | SD | Units |
| Age | 56.19 | 8.34 | 55.83 | 8.26 | 56.61 | 8.41 | Years |
| Body mass index | 27.27 | 4.69 | 26.85 | 5.09 | 27.76 | 4.11 | kg/m$^2$ |
| Glucose | 5.10 | 1.99 | 5.04 | 1.05 | 5.17 | 1.34 | mmol/L |
| HbA1c | 35.81 | 6.44 | 35.48 | 5.80 | 36.20 | 7.09 | mmol/mol |
| Triglycerides | 1.01 | 1.73 | 1.53 | 0.86 | 1.96 | 1.13 | mmol/L |
| Type 2 diabetes | 2,618 cases | - | 865 cases | - | 1753 cases | - | Yes/No. |
| Sample storage time | 665.40 | 43.69 | 665.60 | 43.47 | 665.30 | 43.95 | Weeks |

*Supplementary Table 6. Serum CD300LG cis-pQTLs*

| SNP | Chromosome | Base position | Effect allele | Other allele | Effect allele frequency | INFO score | Effect | SE | *p*-value | F-statistic |
|---|---|---|---|---|---|---|---|---|---|---|
| rs72831096 | 17 | 41469246 | G | A | 0.7937 | 0.9978 | 0.0312 | 0.0033 | $6.3\times10^{-21}$ | 88.0903 |
| rs851057 | 17 | 41837264 | G | C | 0.1283 | 0.9681 | -0.0981 | 0.0041 | $3.8\times10^{-129}$ | 584.5963 |
| rs231474 | 17 | 42016477 | T | G | 0.4528 | 0.9942 | 0.0327 | 0.0027 | $1.1\times10^{-33}$ | 146.4119 |

SNP = single nucleotide polymorphism. SE = standard error.

*Supplementary Table 7. Serum CD300LG cis-pQTLs*

| SNP | CHR | Base position | Effect allele | Other allele | Effect allele frequency | INFO score | Effect | SE | *p*-value |
|---|---|---|---|---|---|---|---|---|---|
| rs76230830 | 1 | 23784293 | A | G | 0.918135 | 0.986397 | 0.027182 | 0.004972 | 4.60E-08 |
| rs4651034 | 1 | 1.79E+08 | A | C | 0.663704 | 0.996121 | -0.01747 | 0.002868 | 1.10E-09 |
| rs3753847 | 1 | 2.05E+08 | T | C | 0.151267 | 0.973821 | 0.021328 | 0.003809 | 2.20E-08 |
| rs12040264 | 1 | 2.2E+08 | G | T | 0.397163 | 0.99713 | 0.016136 | 0.002766 | 5.40E-09 |
| rs1260326 | 2 | 27730940 | T | C | 0.391047 | 1 | -0.02078 | 0.002772 | 6.70E-14 |
| rs75166367 | 2 | 1.63E+08 | G | A | 0.93965 | 1 | 0.03759 | 0.005696 | 4.10E-11 |
| rs12328675 | 2 | 1.66E+08 | T | C | 0.881422 | 0.99872 | -0.02457 | 0.004186 | 4.30E-09 |
| rs56256300 | 2 | 2.27E+08 | A | AT | 0.35506 | 0.999036 | 0.017272 | 0.002841 | 1.20E-09 |
| rs1899951 | 3 | 12394840 | C | T | 0.877417 | 0.997266 | -0.0388 | 0.004135 | 6.30E-21 |
| rs77572134 | 3 | 52895934 | T | C | 0.858413 | 0.997026 | -0.0235 | 0.003886 | 1.50E-09 |
| rs11943206 | 4 | 56437010 | T | C | 0.700938 | 0.99321 | -0.01638 | 0.002954 | 2.90E-08 |
| rs13127398 | 4 | 1.03E+08 | T | A | 0.92883 | 0.985517 | -0.03515 | 0.005331 | 4.30E-11 |
| rs574745983 | 5 | 53302258 | A | AT | 0.752631 | 0.981441 | 0.017945 | 0.003171 | 1.50E-08 |
| rs164514 | 5 | 1.41E+08 | A | G | 0.658169 | 0.996885 | 0.016234 | 0.002874 | 1.60E-08 |
| rs1064627 | 6 | 30698541 | A | G | 0.8 | 0.999598 | -0.01976 | 0.003383 | 5.20E-09 |
| rs614008 | 6 | 31840794 | C | T | 0.421165 | 0.996933 | 0.017261 | 0.002743 | 3.10E-10 |
| rs75104038 | 6 | 34190104 | G | A | 0.938486 | 0.992843 | 0.031943 | 0.005663 | 1.70E-08 |
| rs998584 | 6 | 43757896 | C | A | 0.520758 | 0.994408 | 0.03954 | 0.002717 | 5.40E-48 |
| rs9402846 | 6 | 1.37E+08 | G | C | 0.410445 | 0.990684 | -0.01628 | 0.002767 | 4.00E-09 |
| rs28588745 | 8 | 10647044 | A | T | 0.792571 | 0.995694 | 0.047092 | 0.003354 | 8.60E-45 |
| rs13267474 | 8 | 23784587 | C | T | 0.590946 | 0.971999 | -0.01559 | 0.002791 | 2.30E-08 |
| rs2980888 | 8 | 1.27E+08 | T | C | 0.300429 | 0.999267 | -0.02082 | 0.002966 | 2.20E-12 |
| rs8176747 | 9 | 1.36E+08 | C | G | 0.937707 | 0.999944 | -0.10105 | 0.005604 | 1.10E-72 |
| rs4418728 | 10 | 94839724 | G | T | 0.549296 | 0.998989 | -0.02406 | 0.002728 | 1.10E-18 |
| rs11231698 | 11 | 63877163 | C | T | 0.945696 | 0.986634 | 0.046951 | 0.006031 | 7.00E-15 |
| rs79719909 | 12 | 20474706 | A | G | 0.794547 | 0.977208 | -0.01872 | 0.003388 | 3.30E-08 |
| rs6489191 | 12 | 1.23E+08 | A | G | 0.415161 | 1 | -0.01658 | 0.002765 | 2.00E-09 |
| rs7956959 | 12 | 1.25E+08 | C | T | 0.880147 | 0.99506 | -0.02507 | 0.004168 | 1.80E-09 |
| rs9513116 | 13 | 29016715 | G | A | 0.679419 | 0.98658 | 0.017647 | 0.002923 | 1.60E-09 |
| rs261290 | 15 | 58678720 | T | C | 0.346128 | 0.996264 | 0.025009 | 0.00287 | 3.00E-18 |
| rs1077834 | 15 | 58723479 | T | C | 0.779062 | 0.995244 | -0.02278 | 0.003279 | 3.80E-12 |
| rs36060036 | 16 | 20361950 | C | T | 0.834888 | 0.994847 | 0.027024 | 0.003673 | 1.90E-13 |
| rs56228609 | 16 | 56987765 | C | T | 0.68325 | 0.995814 | -0.02783 | 0.002932 | 2.30E-21 |
| rs2925979 | 16 | 81534790 | T | C | 0.301454 | 1 | -0.02147 | 0.002973 | 5.20E-13 |
| rs2229611 | 17 | 41063466 | T | C | 0.214629 | 1 | -0.02415 | 0.003276 | 1.70E-13 |
| 17:42300634_CAAAAA_C | 17 | 42300634 | CAAAAA | C | 0.616177 | 0.982143 | -0.02467 | 0.002791 | 9.70E-19 |
| rs2306828 | 17 | 43213772 | A | C | 0.164834 | 0.988568 | -0.02401 | 0.003651 | 4.90E-11 |
| rs4969142 | 17 | 76398304 | G | A | 0.510431 | 0.99707 | -0.01551 | 0.002703 | 9.50E-09 |
| rs7412 | 19 | 45412079 | C | T | 0.917544 | 1 | -0.03758 | 0.004889 | 1.50E-14 |
| rs6088669 | 20 | 30177582 | C | A | 0.843559 | 0.997585 | -0.02721 | 0.003746 | 3.70E-13 |

SNP = single nucleotide polymorphism. CHR = chromosome. SE = standard error.

*Supplementary Table 8. Results of cis-only pQTLs MR analysis for serum protein level (log2) on outcomes of interest (2-hour post OGTT glucose (mmol/L), fasting glucose(mmol/L), fasting insulin(mmol/L), and HbA1c (%)).*

| Outcome | N SNPs | Effect estimate | SE | p-value | Q heterogeneity | Q p-value |
|---|---|---|---|---|---|---|
| 2-hour post OGTT glucose | 3 | -0.1285 | 0.1150 | 0.2640 | 0.9401 | 0.6250 |
| Fasting glucose | 3 | -0.0203 | 0.0346 | 0.5580 | 3.8401 | 0.1466 |
| Fasting insulin | 3 | -0.0681 | 0.0277 | 0.0139 | 0.9232 | 0.6303 |
| HbA1c | 3 | -0.0357 | 0.0319 | 0.2626 | 6.1030 | 0.0473 |

*Supplementary Table 9. Results of cis- and trans-combined pQTLs MR analysis for outcomes of interest (2-hour post OGTT glucose, fasting glucose, fasting insulin, and HbA1c).*

| Outcome | Method | N SNPs | Effect estimate | SE | *p*-value | Q heterogeneity | Q p-value | Intercept | Intercept p-value |
|---|---|---|---|---|---|---|---|---|---|
| 2-hour post OGTT glucose | IVW | 39 | -0.3722 | 0.0998 | $1.92\times10^{-4}$ | 128.8568 | $8.22\times10^{-12}$ | | |
| 2-hour post OGTT glucose | MR Egger | 39 | -0.0455 | 0.1957 | 0.8174 | | | -0.0115 | 0.0628 |
| 2-hour post OGTT glucose | Weighted median | 39 | -0.1163 | 0.0916 | 0.2039 | | | | |
| 2-hour post OGTT glucose | Simple mode | 39 | -0.2194 | 0.1702 | 0.2051 | | | | |
| 2-hour post OGTT glucose | Weighted mode | 39 | -0.1241 | 0.0831 | 0.1436 | | | | |
| Fasting glucose | IVW | 39 | -0.0307 | 0.0358 | 0.3909 | 349.5152 | $5.59\times10^{-52}$ | | |
| Fasting glucose | MR Egger | 39 | -0.0327 | 0.0749 | 0.6645 | | | $6.89\times10^{-5}$ | 0.9758 |
| Fasting glucose | Weighted median | 39 | -0.0292 | 0.0202 | 0.1488 | | | | |
| Fasting glucose | Simple mode | 39 | -0.1040 | 0.0385 | 0.0102 | | | | |
| Fasting glucose | Weighted mode | 39 | -0.0175 | 0.0221 | 0.4342 | | | | |
| Fasting insulin | IVW | 39 | -0.0870 | 0.0559 | 0.1188 | 666.0874 | $9.63\times10^{-116}$ | | |
| Fasting insulin | MR Egger | 39 | 0.0354 | 0.1126 | 0.7547 | | | -0.0043 | 0.2195 |
| Fasting insulin | Weighted median | 39 | -0.0657 | 0.0255 | 0.0100 | | | | |
| Fasting insulin | Simple mode | 39 | -0.0280 | 0.0428 | 0.5164 | | | | |
| Fasting insulin | Weighted mode | 39 | -0.0347 | 0.0328 | 0.2972 | | | | |
| HbA1c | IVW | 39 | -0.0485 | 0.0155 | 0.0017 | 115.4975 | $9.53\times10^{-10}$ | | |
| HbA1c | MR Egger | 39 | -0.0610 | 0.0319 | 0.0638 | | | 0.0004 | 0.6565 |
| HbA1c | Weighted median | 39 | -0.0513 | 0.0159 | 0.0013 | | | | |
| HbA1c | Simple mode | 39 | -0.0522 | 0.0344 | 0.1380 | | | | |
| HbA1c | Weighted mode | 39 | -0.0465 | 0.0213 | 0.0351 | | | | |

*Supplementary Table 10: SNPs and their prior associations*

| SNP | CHR | BP | ALLELE1 | ALLELE0 | A1FREQ | BETA | SE | P | Phenoscanner |
|---|---|---|---|---|---|---|---|---|---|
| rs36060036 | 16 | 20361950 | C | T | 0.835 | 0.027 | 0.004 | 1.9E-13 | Blood pressure, |
| rs6088669 | 20 | 30177582 | C | A | 0.844 | -0.027 | 0.004 | 3.7E-13 | Blood pressure, |
| rs28588745 | 8 | 10647044 | A | T | 0.793 | 0.047 | 0.003 | 8.2E-45 | Body fat percentage |
| rs2306828 | 17 | 43213772 | A | C | 0.165 | -0.024 | 0.004 | 4.9E-11 | Body mass, |
| rs614008 | 6 | 31840794 | C | T | 0.421 | 0.017 | 0.003 | 3.0E-10 | Body mass, Hip circumference, blood pressure |
| rs75104038 | 6 | 34190104 | G | A | 0.938 | 0.032 | 0.006 | 1.7E-08 | Body mass, Waist circumference |
| rs9513116 | 13 | 29016715 | G | A | 0.679 | 0.018 | 0.003 | 1.5E-09 | CAD |
| rs56228609 | 16 | 56987765 | C | T | 0.683 | -0.028 | 0.003 | 2.3E-21 | CAD, HDL, |
| rs261290 | 15 | 58678720 | T | C | 0.346 | 0.025 | 0.003 | 3.0E-18 | Cholesterol |
| rs7412 | 19 | 45412079 | C | T | 0.918 | -0.038 | 0.005 | 1.5E-14 | Cholesterol, APOB, CAD, Weight, MI, Waist circumference, BMI |
| rs1077834 | 15 | 58723479 | T | C | 0.779 | -0.023 | 0.003 | 3.7E-12 | Cholesterol, Triglycerides, |
| rs8176747 | 9 | 136131315 | C | G | 0.938 | -0.101 | 0.006 | 1.1E-72 | Chronic kidney disease |
| rs1899951 | 3 | 12394840 | C | T | 0.877 | -0.039 | 0.004 | 6.3E-21 | Fat mass, diabetes type 2, BMI, WHR, insulin sensitivity |
| rs1064627 | 6 | 30698541 | A | G | 0.800 | -0.020 | 0.003 | 5.1E-09 | Fat mass/fat-free mass, blood pressure, hip circumfrence, cholesterol, diabetes |
| rs231492 | 17 | 41978756 | G | T | 0.916 | 0.155 | 0.005 | 4.1E-218 | HDL |
| rs2925979 | 16 | 81534790 | T | C | 0.301 | -0.021 | 0.003 | 5.2E-13 | HDL, WHR, Diabetes type 2, triglycerides, BMI |
| rs11231698 | 11 | 63877163 | C | T | 0.946 | 0.047 | 0.006 | 7.0E-15 | Hypertention |
| rs4418728 | 10 | 94839724 | G | T | 0.549 | -0.024 | 0.003 | 1.1E-18 | Triglycerides, body fat percentage |
| rs1260326 | 2 | 27730940 | T | C | 0.391 | -0.021 | 0.003 | 6.5E-14 | Triglycerides, cholesterol, CRP, glucose, fat mass, alcohol intake, weight and more |
| rs2980888 | 8 | 126507308 | T | C | 0.300 | -0.021 | 0.003 | 2.3E-12 | Triglycerides, cholesterol, fat percentage, hip circumference, CAD |
| rs998584 | 6 | 43757896 | C | A | 0.521 | 0.040 | 0.003 | 5.3E-48 | WHR, Hip circumference, Triglycerides, Cholesterol, BMI |
